# *IFIH1* rs1990760 variants, systemic inflammation and outcome in critically-ill COVID-19 patients

**DOI:** 10.1101/2021.07.03.21259946

**Authors:** Laura Amado-Rodríguez, Estefanía Salgado del Riego, Juan Gómez de Oña, Inés López-Alonso, Helena Gil-Peña, Cecilia López-Martínez, Paula Martín-Vicente, Antonio López-Vázquez, Adrián González-López, Elías Cuesta-Llavona, Raquel Rodríguez-García, José Antonio Boga, Marta Elena Álvarez-Arguelles, Juan Mayordomo-Colunga, José Ramón Vidal-Castiñeira, Irene Crespo, Margarita Fernández, Loreto Criado, Victoria Salvadores, Francisco José Jimeno-Demuth, Lluís Blanch, Belén Prieto, Alejandra Fernández-Fernández, Carlos López-Larrea, Eliecer Coto, Guillermo M Albaiceta

## Abstract

**Rationale:** Outcomes in patients with severe SARS-CoV-2 infection (COVID-19) are conditioned by virus clearance and regulation of inflammation. Variants in *IFIH1*, a gene coding the cytoplasmatic RNA sensor MDA5, regulate the response to viral infections.

**Objective:** To characterize the impact of *IFIH1* rs199076 variants on host response and outcomes after severe COVID-19.

**Methods:** Patients admitted to an intensive care unit (ICU) with confirmed COVID-19 were prospectively studied and rs1990760 variants determined. Peripheral blood gene expression, cell populations and immune mediators were measured. Peripheral blood mononuclear cells from healthy volunteers were exposed to an MDA5 agonist and dexamethasone *ex-vivo*, and changes in gene expression assessed. ICU discharge and hospital death were modelled using rs1990760 variants and dexamethasone as factors in this cohort and *in-silico* clinical trials.

**Measurements and Main Results:** 227 patients were studied. Patients with the *IFIH1* rs1990760 TT variant showed a lower expression of inflammation-related pathways, an anti-inflammatory cell profile and lower concentrations of pro-inflammatory mediators. Cells with TT variant exposed to a MDA5 agonist showed an increase in *IL6* expression after dexamethasone treatment. All patients with the TT variant not treated with steroids (N=14) survived their ICU stay (HR 2.49, 95% confidence interval 1.29–4.79). Patients with a TT variant treated with dexamethasone (N=50) showed an increased hospital mortality (HR 2.19, 95% confidence interval 1.01–4.87) and serum IL-6. *In-silico* clinical trials supported these findings.

**Conclusions:** COVID-19 patients with the *IFIH1* rs1990760 TT variant show an attenuated inflammatory response and better outcomes. Dexamethasone may reverse this anti-inflammatory phenotype.

## Introduction

The spectrum of disease after infection by SARS-CoV-2 (COVID-19) may range from mild respiratory symptoms to a severe form of lung injury fulfilling acute respiratory distress syndrome (ARDS) criteria (1). In these severe cases, systemic response to infection may be associated to multiorgan failure and death (2). Therefore, outcomes in critically ill COVID-19 patients are related not only to viral clearance, but also to preservation of homeostasis.

The mechanisms responsible for the development of severe forms of COVID-19 have not been fully elucidated, but non-adaptative inflammatory responses play a central role. The only strategies that have decreased mortality in this population, steroids (3) and blockade of the IL-6 pathway (4), aim to limit this exacerbated immune response to prevent organ dysfunction.

The human gene *IFIH1*, located in the reverse strand of chromosome 2, encodes MDA5, a helicase that acts as a cytoplasmatic virus receptor. After binding to a viral RNA strand, MDA5 interacts with a mitochondrial adapter (MAVS, mitochondrial antiviral signaling protein), triggering the transcription of type-1 interferon genes and ultimately the systemic inflammatory response. In humans, the rs1990760 polymorphism encodes a variant of the *IFIH1* gene (NM_022168: c.2836G>A [p.Ala946Thr]) that has been related to different susceptibility to viral infections and autoimmune disorders (5). By regulation of IFN-dependent pathways, *IFIH1* participates in a feedback loop that ultimately modulates viral clearance and host inflammatory responses. In experimental models of Coxsackie virus infection, TT variants in rs1990760 result in lower proinflammatory cytokine levels without a major reduction in viral clearance (6).

Although the role of this variant in Coronavirus infections has not been explored, it has been proposed that the rs1990760 TT variant could confer resistance to SARS-CoV-2 infection and that differences in allelic frequencies could explain the epidemiological features of the pandemic in different countries (7). We hypothesized that, once infection is established, the inflammatory response in severe COVID-19 patients could be conditioned by *IFIH1* variants. To test this hypothesis, we prospectively followed a cohort of critically ill patients with confirmed infection by SARS-CoV-2, in which peripheral blood gene expression, cell populations, concentrations of immune mediators and clinical outcomes were studied and related to rs1990760 variants.

## Methods

A detailed description of the methods is provided in the online supplement.

### Study design

This single-center prospective, observational study was approved (ref. 2020/188) by the Clinical Research Ethics Committee of Principado de Asturias (Spain). Informed consent was obtained from each participant or next of kin. Given the exploratory nature of the study objective and the absence of previous data, no formal sample size calculations were performed.

All patients with confirmed SARS-CoV-2 infection (8) and meeting the Kigali modification (9) of ARDS criteria (to include patients without mechanical ventilation) from 16-Mar-2020 to 10-Dec-2020 were included in the study. Exclusion criteria were age<18, any condition that could explain the respiratory failure other than COVID-19, do-not-resuscitate orders or terminal status, or refusal to participate. The presence of SARS-CoV-2 in respiratory samples was detected by reverse transcriptase qPCR (Supplementary table E1) and clearance half-life (λ_Viral Clearance_) calculated (10).

Patients were followed until hospital discharge, and clinical and analytical data collected. The main outcome was ICU discharge alive and spontaneously breathing. Secondary outcome was hospital discharge.

### Biochemical measurements

*IFIH1* rs1990760 C/T polymorphism was determined by PCR genotyping in DNA extracted from peripheral blood. Blood samples were taken in days 1 (first 24 hours after ICU admission, before steroid treatment) and 4 of ICU stay. RNA was extracted and sequenced. The generated sequences were mapped and counted using Salmon v1.4 software (11). Raw counts were compared between groups using DESeq2 (12). Genes with differential expression (adjusted p-value<0.05, corrected using a false discovery rate of 0.05) were analyzed using Ingenuity Pathway Analysis (Qiagen, USA) to identify overrepresented gene sets and networks. Over the identified networks, *in-silico* effects of *IFIH1* down-regulation and addition of exogenous dexamethasone were tested. Circulating cell populations were estimated from gene expression using a previously validated deconvolution algorithm (13). The correlation between estimated and measured lymphocyte percentages was 0.61 (Supplementary Figure E1).

In a second sample, a panel of serum inflammatory mediators (interferons (IFN)-β, -γ and -λ, tumor necrosis factor (TNF)-α, interleukins (IL)-1β and -6, and chemokines CXCL8, CXCL9, CXCL10, CXCL16, CCL2, CCL3, CCL4 and CCL7) was measured using a multiplexed assay. Additional serum IL-6 measurements were done during clinical follow-up.

### Ex-vivo experiments

Peripheral blood mononuclear cells (PBMC) from healthy volunteers, genotyped for *IFIH1* rs1990760, were isolated and cultured in presence of medium, medium plus a MDA5 ligand (high-molecular weight poly-I:C), or medium plus the MDA5 ligand and dexamethasone (Kern Pharma, Spain). After 24h cells were collected and homogenized, RNA extracted and expression of *STAT1, STAT3, FOXO3*, I*L6* and *GAPDH* quantified (Supplementary Table E2).

### Clinical trial simulations

Data from the RECOVERY trial were extracted and used to estimate risk ratios (RR) for each rs1990760 variant, with and without steroids (see online supplement for details). With these data, a survival model was developed and used to simulate scenarios with different allelic frequencies and baseline risks of death. Hazard ratios (HR) for 28-day mortality were calculated from these simulations.

### Statistical analysis

Data are shown as median (interquartile range). Comparisons between *IFIH1* variants were done using Wilcoxon or ANOVA tests, and p-values corrected using a false discovery rate of 0.05. Results from the *ex-vivo* experiments were fitted to a mixed-effects model including donor as a fixed effect and experimental group and genotype as random effects. Survival was analyzed using competing risks models considering ICU/hospital discharge alive and spontaneously breathing and ICU/hospital death as different terminal events. *IFIH1* variants and steroid treatment were added in these models as covariables with an interaction, thus resulting in 4 study groups. Patients with rs1990760 CC/CT variants not treated with steroids were considered the reference category in all the analyses. All the analyses and plots were performed using R 4.0.1 (14).

## Results

### Study cohort

250 patients were admitted in ICU due to suspected or confirmed COVID-19. Among these, 227 were included in the study. The study flow chart and reasons for exclusion are shown in Figure 1. Basic demographic and clinical data are shown in table 1. *IFIH1* rs1990760 variants in this population met Hardy-Weinberg conditions (53 CC, 110 CT, 64 TT, chi-square p=0.19). There were no differences in comorbidities or clinical data at admission between genotypes other than a lower PaO_2_/FiO_2_ ratio in patients with a TT variant (Table 1).

**Table 1.** Clinical characteristics of the study cohort. COPD: Chronic Obstructive Pulmonary Disease. Values are shown as absolute count or median (interquartile range). PBW: Predicted body weight. NIV: Non-invasive ventilation. PEEP: Positive End-Expiratory Pressure. *p values calculated for proportion over the number of intubated patients.

|  | Overall (n=227) | rs1990760 |  | P value |
| --- | --- | --- | --- | --- |
|  |  | TT (n=65) | CC/CT (n=162) |  |
| Age (y) | 67 (60 - 75) | 68.5 (63 - 75.25) | 66 (58 - 75) | 0.123 |
| Sex |  |  |  | 0.327 |
| Male | 174 (77) | 47 (72) | 127 (78) |  |
| Female | 53 (23) | 18 (28) | 35 (22) |  |
| Race |  |  |  | 0.527 |
| Black | 3 (1.5) | 1 (2) | 2 (1.5) |  |
| White | 207 (91) | 62 (95) | 146 (90) |  |
| Latino | 15 (7) | 2 (3) | 13 (8) |  |
| Asian | 1 (0.5) | 0 | 1 (0.5) |  |
| Body mass index (Kg/m<sup>2</sup>) | 30 (27 - 33) | 30 (26 - 33) | 30 (27 - 32) | 0.592 |
| Days since symptom onset | 8 (6 - 11) | 8 (6 - 10) | 8 (6 - 11) | 0.873 |
| Days from hospital admission to ICU admission | 2 (1-4) | 2 (1 - 3) | 2 (1 - 4) | 0.841 |
| Arterial hypertension | 132 (58) | 38 (59) | 94 (58) | 0.932 |
| Diabetes | 51 (23) | 14 (22) | 37 (23) | 1 |
| Chronic kidney disease | 15 (7) | 5 (8) | 10 (6) | 0.743 |
| COPD | 17 (8) | 7 (11) | 10 (6) | 0.339 |
| Cirrhosis | 2 (1) | 1 (2) | 1 (0.5) | 1 |
| Neoplasms |  |  |  | 0.669 |
| No | 215 (95) | 62 (95) | 154 (95) |  |
| Active | 10 (4) | 3 (5) | 7 (3.5)) |  |
| Past | 2 (1) | 0 | 2 (1.5) |  |
| Immunosuppressive drugs (incl. steroids) | 5 (2) | 3 (5) | 2 (1.5) | 0.142 |
| Ventilation at admission |  |  |  | 0.278 |
| Spontaneous /NIV | 38 (17) | 11 (17) | 27 (17) |  |
| Controlled invasive ventilation | 187 (82.5) | 53 (82) | 135 (83) |  |
| Pressure support ventilation | 1 (0.5) | 1 (2) | 0 |  |
| FiO<sub>2</sub> | 0.5 (0.4 - 0.6) | 0.5 (0.4 - 0.7) | 0.5 (0.4 - 0.6) | 0.104 |
| PaO<sub>2</sub>/FiO<sub>2</sub> (mmHg) | 204 (155 - 267) | 185 (142 - 226) | 218 (162 - 282) | 0.007 |
| PaCO<sub>2</sub> (mmHg) | 43 (39 - 47) | 44 (39- 47) | 43 (39 - 48) | 0.924 |
| Respiratory rate (min<sup>-1</sup>) | 18 (16 - 20) | 18 (16 - 20) | 18 (16 - 22) | 0.929 |
| Arterial pH | 7.38<br>(7.32 - 7.41) | 7.38<br>(7.33 - 7.42) | 7.37<br>(7.32 - 7.41) | 0.805 |
| Tidal volume/PBW (ml/Kg) | 7.6 (6.9 - 8.4) | 8 (6.9 - 9.2) | 7.5 (7 - 8.1) | 0.109 |
| Plateau pressure (cmH<sub>2</sub>O) | 24 (21 - 28) | 24 (22 - 29) | 24 (21 - 27) | 0.221 |
| PEEP (cmH<sub>2</sub>O) | 12 (10 - 14) | 12 (10 - 14) | 12 (10 - 14) | 0.397 |
| Driving pressure (cmH<sub>2</sub>O) | 12 (10 - 14) | 12 (10 - 14) | 12 (10 - 14) | 0.496 |
| Respiratory system compliance | 38 (32 - 50) | 36 (31 - 44) | 42 (32 - 50) | 0.11 |

(ml/cmH<sub>2</sub>O)
| Leukocytes (x10<sup>3</sup>/μl) | 8.34<br>(6.05 – 11.38) | 8.77<br>(5.96 – 11.52) | 8.29<br>(6.08 – 11.18) | 0.773 |
| --- | --- | --- | --- | --- |
| Lymphocytes (x10<sup>3</sup>/μl) | 0.65<br>(0.48 – 0.93) | 0.62<br>(0.47 – 0.94) | 0.67<br>(0.49 – 0.93) | 0.266 |
| Serum creatinine (mg/dl) | 0.83<br>(0.62 - 1.13) | 0.87<br>(0.62 - 1.14) | 0.82<br>(0.62 - 1.11) | 0.859 |
| Serum ferritin (ng/ml) | 1107<br>(717 - 1667) | 965<br>(634 - 1263) | 1262<br>(776 - 2374) | 0.011 |
| D-dimer (ng/ml) | 1053<br>(681 - 1967) | 1085<br>(643 - 1805) | 1036<br>(683 - 2126) | 0.854 |
| Steroids | 178 (78) | 51 (79) | 127 (78) | 1 |
| Vasoactive drugs | 102 (45) | 27 (42) | 75 (46) | 0.686 |
| Invasive mechanical ventilation | 210 (93) | 59 (91) | 151 (93) | 0.724 |
| Neuromuscular blocking agents | 99 (44) | 30 (46) | 69 (43) | 0.733* |
| Prone ventilation | 119 (52) | 32 (49) | 87 (54) | 0.657* |
| ECMO | 5 (2) | 2 (3) | 3 (2) | 0.544* |

**Figure 1.**
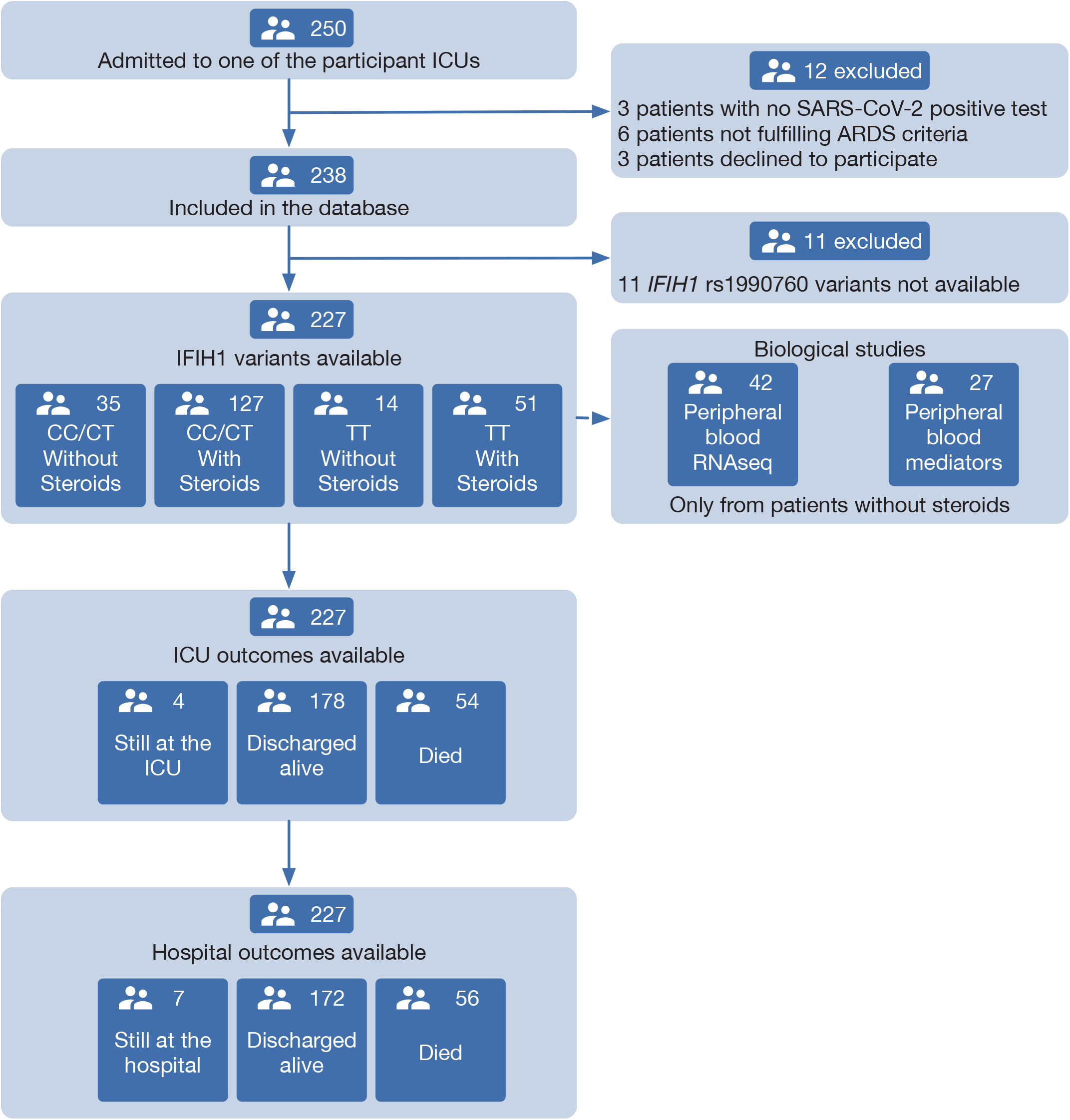
Patient study flow chart. From 250 patients admitted to the participant ICUs during the study period, 227 were included in the study and classified according to the *IFIH1* rs1990760 variants and prescription of steroid therapy. Peripheral blood samples for gene expression (N=41) and quantification of immune mediators (N=28) were taken in the first day after ICU admission in patients not receiving steroids. All patients were followed up until death or hospital discharge.

### IFIH1 rs1190760 variants and inflammatory response to SARS-CoV-2

Gene expression in peripheral blood during the first day of ICU admission was profiled in 42 patients who did not receive steroid therapy at that time (11, 19 and 12 with rs1990760 TT, CT and CC genotypes respectively). Expression of *IFIH1* was significantly lower in patients with the TT genotype (Figure 2A and Supplementary Figure E2). Comparison of peripheral blood gene expression between patients with TT and CT/CC variants yielded significant differences in 179 genes (Figure 2B-C, Supplementary File 1). Visual inspection of the heatmap reveals that differences are quantitative rather than qualitative. Ingenuity pathway analysis revealed several gene networks involved in the regulation of the inflammatory response among the differentially expressed genes (Figure 2D and Supplementary Figure E3). *In-silico* predictions suggests that *IFIH1* downregulation (such as in patients with the rs1990760 TT variant) decreases expression of proinflammatory pathways (Supplementary Figure E4).

**Figure 2.**
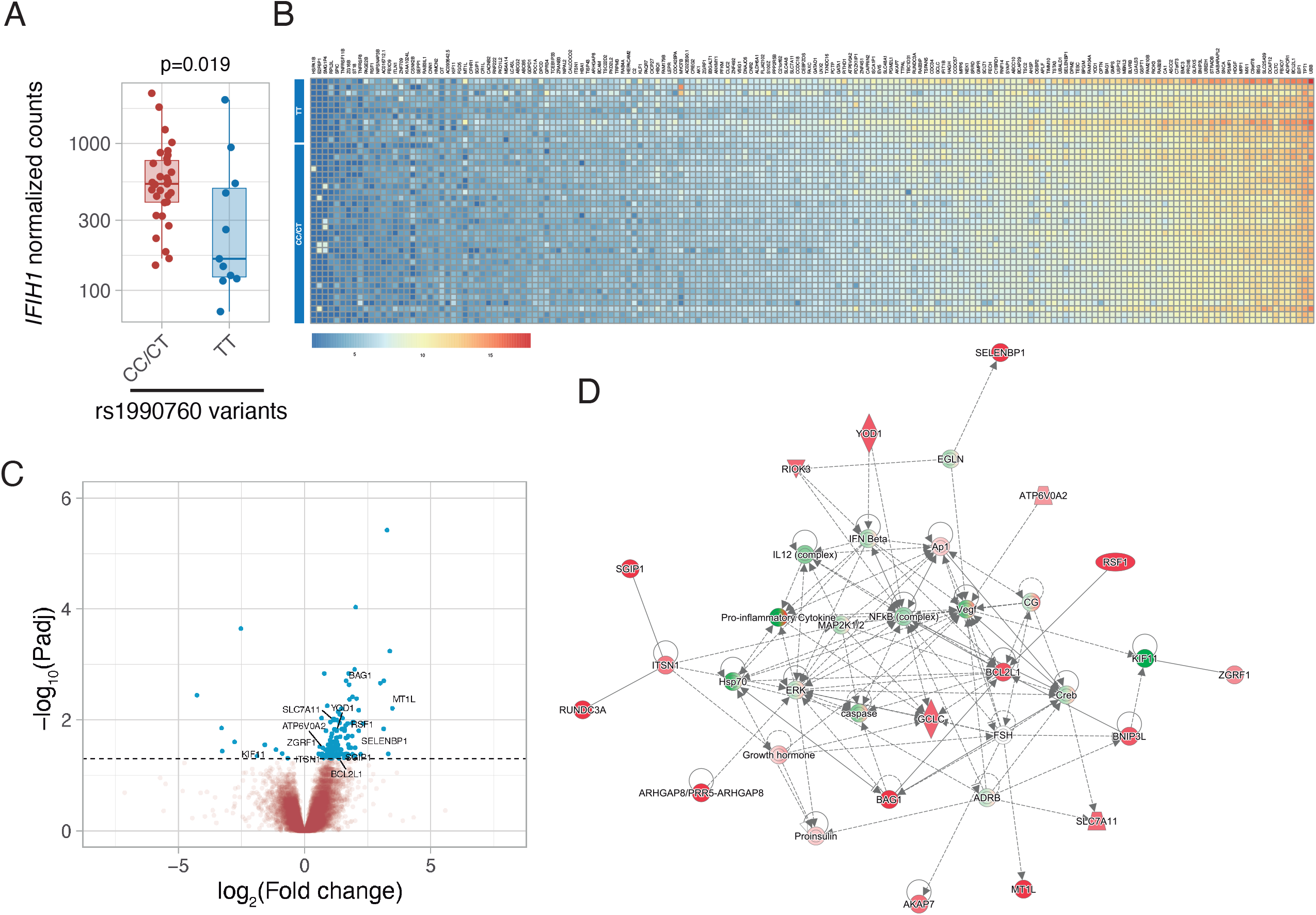
Differences in gene expression according to *IFIH1* rs1990760 genotypes. A: Expression of IFIH1 in peripheral blood in patients with CC/CT (n=31) and TT (n=11) variants (p value calculated using a Wilcoxon test). B: Heatmap showing expression of the 160 genes with significant differences between variants. C: Volcano plot showing the distribution of the magnitude of the differences in gene expression (Log2 Fold change) and their statistical significance. Inflammation-related genes with differential expression and included in the network shown in Panel D are labelled. D: Inflammation-related gene network identified using Ingenuity Pathway Analysis on the RNAseq data.

Then we assessed the immunological consequences of these differences in gene expression and cell populations. Patients with the TT genotype have lower inferred percentages of classic CD14+ monocytes and circulating plasma cells, and an increase in hematopoietic precursors, myeloid dendritic cells, M2 macrophages and CD56dim NK cells (Figure 3).

**Figure 3.**
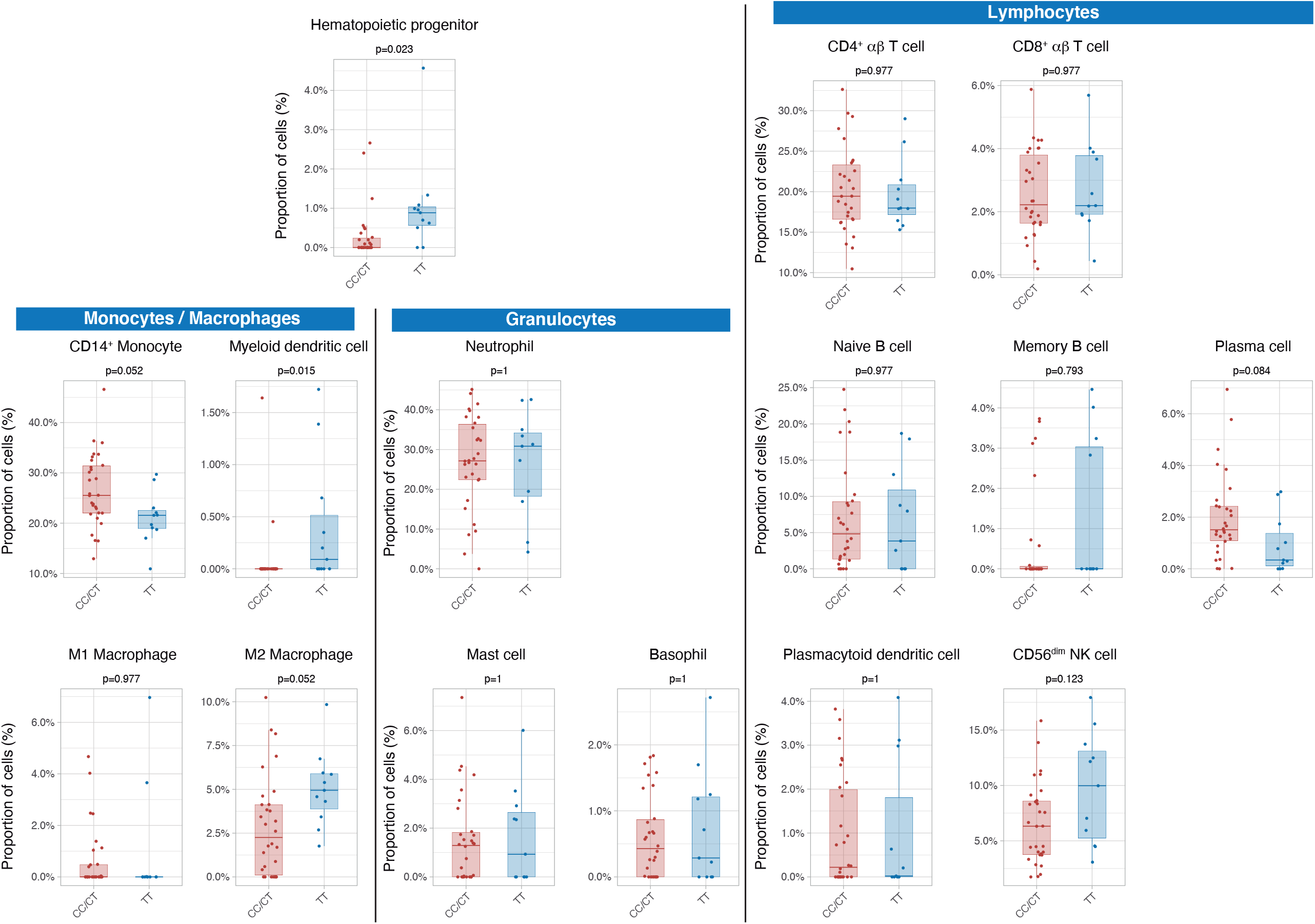
Differences in circulating cell populations according to *IFIH1* rs1990760 variants (31 and 11 samples from patients with a CC/CT or TT variant respectively). Proportions of each cell line were estimated by deconvolution of RNAseq data. P values were calculated using a Wilcoxon test and adjusted using the Benjamini-Hochberg method for a false discovery rate of 5%.

Serum cytokines were measured at ICU admission in 28 patients (8, 10 and 10 with TT, CT and CC genotypes respectively). There were no differences in any of the measured interferons. Patients with the TT genotype showed lower levels of proinflammatory mediators including IL-6, CXCL10, CXCL16, and CCL7 (Figure 4).

**Figure 4.**
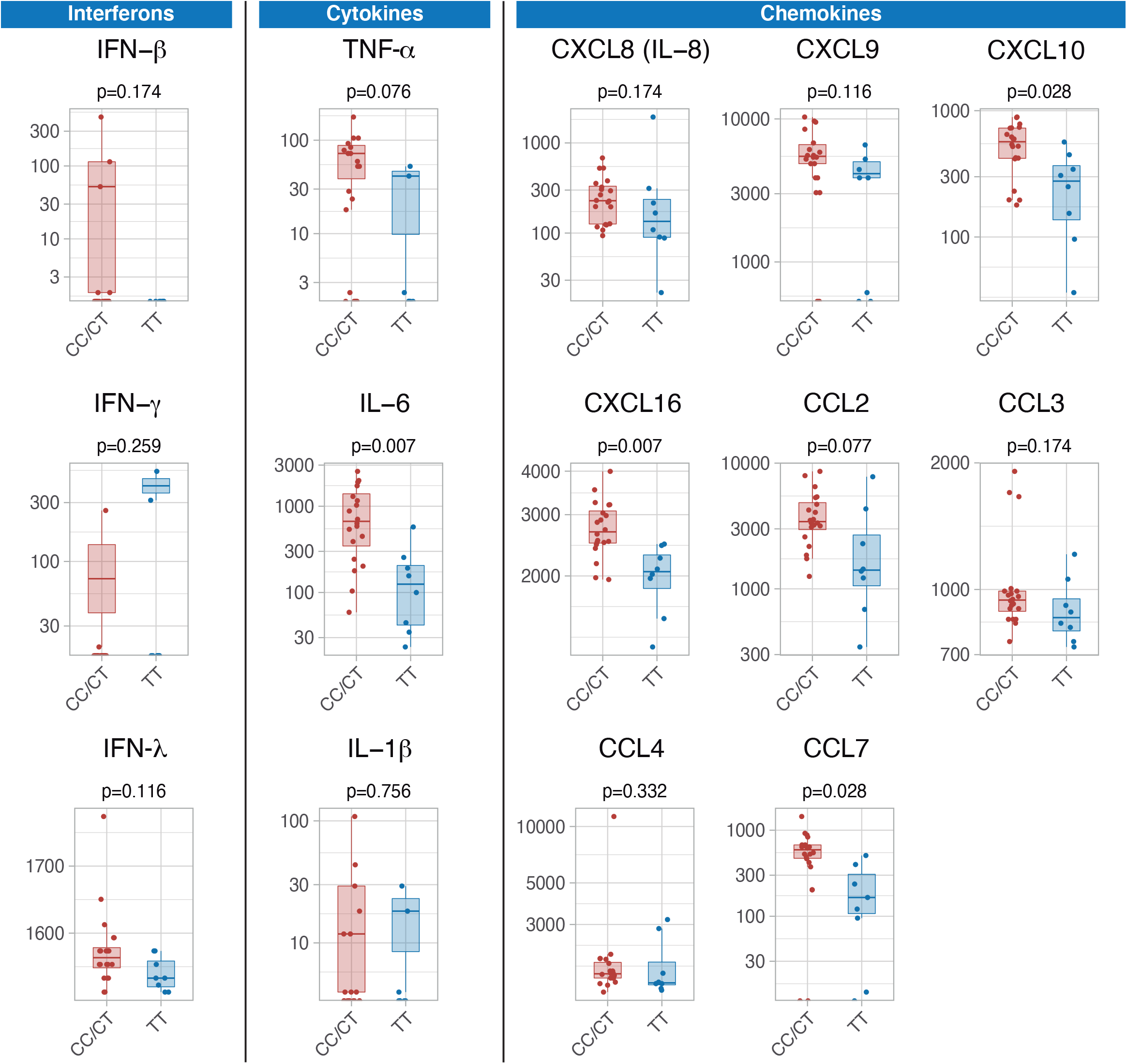
Differences in serum immune mediators according to *IFIH1* rs1990760 variants (19 and 8 samples from patients with a CC/CT or TT variant respectively). P values were calculated using a Wilcoxon test and adjusted using the Benjamini-Hochberg method for a false discovery rate of 5%.

### Ex-vivo effects of dexamethasone in IFIH1 variants

During the study period, dexamethasone was added to COVID-19 treatment based on results from published trials (3). An *in-silico* analysis focused on the interactions between *IFIH1* and steroids suggested that dexamethasone could disrupt some of the effects caused by *IFIH1* downregulation (Supplementary Figure E5). Specifically, dexamethasone may change expression of several *IFIH1*-dependent genes, including *STAT1, STAT3* or *FOXO3* among others.

To test these predictions, an *ex-vivo* experiment using peripheral blood mononuclear cells from healthy volunteers with different *IFIH1* rs1990760 variants (n=5, 6 and 7 for CC, CT and TT variants respectively) was performed. Cells were collected and exposed to poly I:C, to mimic SARS-Cov-2 infection, and dexamethasone. Compared to exposure to poly I:C alone, dexamethasone had no effect on *STAT1* (Figure 5A) or *STAT3* (Figure 5B) expression. However, the steroid increased the expression of *FOXO3* (Figure 5C) and *IL6* (Figure 5D) only in cells with the TT variant. These results suggest that dexamethasone may alter the inflammatory response triggered by MDA5 activation in those patients with the TT variant of the gene.

**Figure 5.**
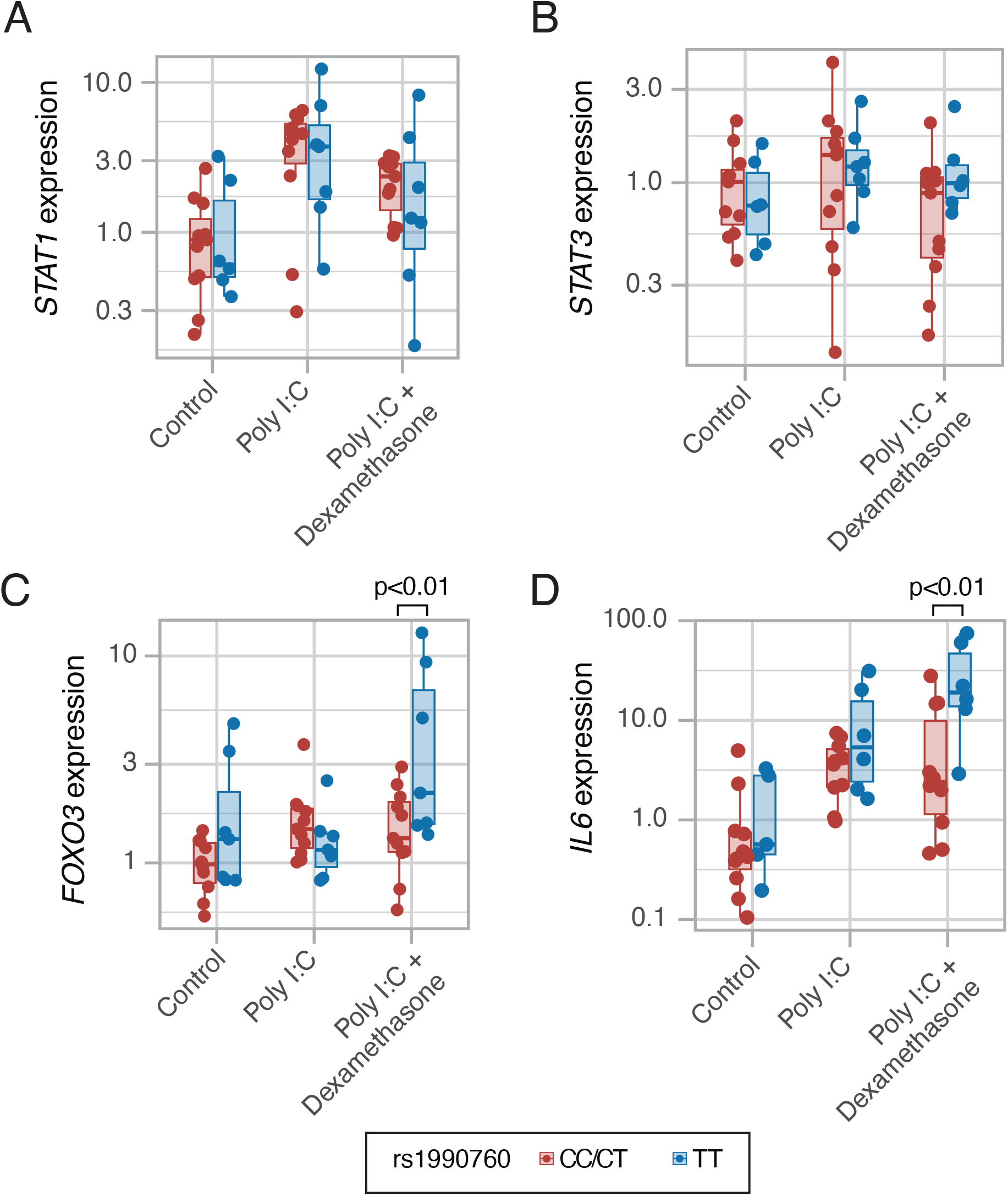
Expression of transcription factors *STAT1* (A, p=0.012, no significant differences between variants in *post-hoc* tests), *STAT3* (B, p=0.443) and *FOXO3* (C, p=0.002) and cytokine *IL6* (D, p<0.001) induced by MDA5 activation using high molecular weight poly-I:C (a viral RNA analogue) and dexamethasone in peripheral mononuclear blood cells. Results were fitted using a mixed-effects linear model including cell donor as a fixed effect and experimental group and genotype as random effects. Pairwise p-values lower than 0.05 (using Holm’s correction) are shown.

### Impact of IFIH1 variants on outcomes in critically ill COVID-19 patients

Median follow-up was 25 days (interquartile range 16 – 39 days) and 27 days in survivors (interquartile range 17 – 44 days). There were no differences in ICU length of stay among groups (Supplementary table E2). ICU survival and hospital mortality were modelled using *IFIH* rs1990760 genotype and steroid treatment as interacting covariables. There were no significant differences in the main clinical characteristics among the resulting groups (Supplementary Table E2). Regarding ICU survival (Figure 6A and Supplementary Table E3), 27 out of 35 patients with CC/CT alleles who did not receive steroids were discharged alive and spontaneously breathing from the ICU (HR 1, used as reference). In patients with this variant, dexamethasone was not related to a better outcome (98 out of 128 patients discharged, HR 1.20 [0.78 – 1.38], p=0.41). All patients with the TT allele (n=14) who did not receive steroids were discharged alive (HR 2.49 [1.29 – 4.79], p=0.012). Steroid treatment in patients with the TT variant was related to the loss of this benefit (32 out of 50 patients discharged alive, HR=1.03 [0.62 – 1.72], p=0.91).

**Figure 6.**
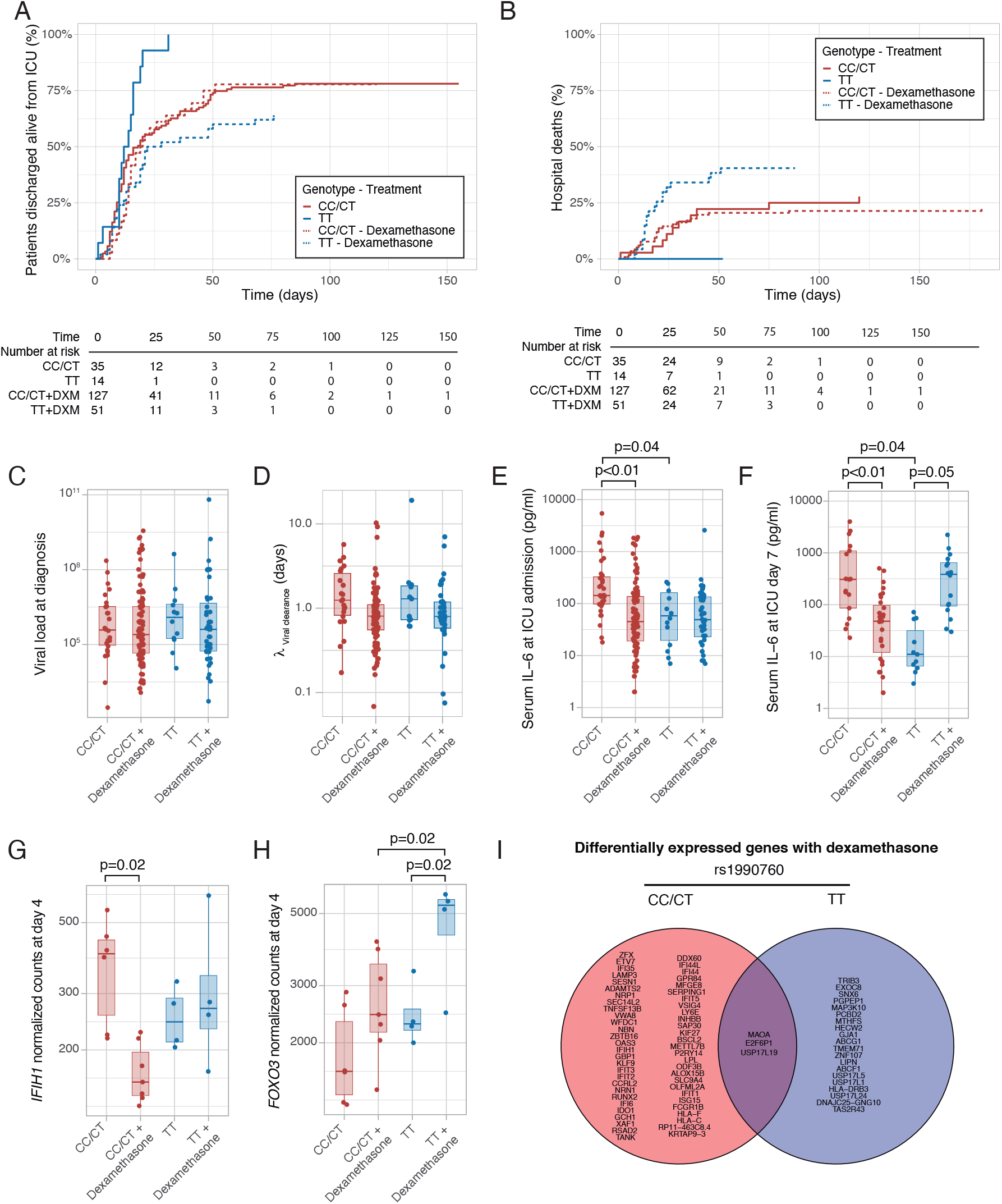
Outcomes according to *IFIH1* rs1990760 variants and treatment with dexamethasone. A: Cumulative incidence of ICU discharge alive and spontaneously breathing for each group. B: Cumulative incidence of hospital death for each group. Outcomes were modelled as competing events. C: Viral load at diagnosis for each group (p=0.888). D: Viral clearance evaluated as half-life of an exponential decay function calculated from the maximal viral load in each patient (p=0.056, no significant differences in post-hoc tests). E-F: Serum IL-6 concentrations during the first day (E, p<0.001) and 7 days (F, p=0.014) after ICU admission for each group. Differences among groups in panels C-F were evaluated using an analysis of the variance. Pairwise p-values lower than 0.05 (using Holm’s correction) are shown. G-H: Changes in *IFIH1* (G) and *FOXO3* (H) expression in day 4 of ICU stay for each group (n=6 and 7 for CC/CT variants without and with dexamethasone respectively, n= 4 and 4 for TT variant without and with dexamethasone respectively). I: Genes with dexamethasone-induced changes in gene expression in ICU day 4 in each rs1990760 variant.

Hospital mortality is shown in Figure 6B and Supplementary Table E3. There were 9 hospital deaths in patients with the CT/CC genotypes not treated with steroids (out of a total of 35 patients), and 27 out of 128 hospital deaths when treated with steroids, resulting in a HR of 1.11 [0.52 – 2.37] (p=0.80). All patients with the TT allele who were not treated with steroids survived after their hospital stay (HR 1.23 × 10^−7^, confidence intervals and p value cannot be computed due to the absence of events). Patients with the TT allele and treated with steroids showed a worse outcome (19 deaths in 50 patients, HR 2.19 [1.01 – 4.87], p=0.05).

To investigate the causes responsible for the differences in mortality among groups, we first quantified viral load at diagnosis (Figure 6C) and clearance after its peak value (Figure 6D). There were no differences in these parameters related to *IFIH1* variants or treatment groups. We also compared serum IL-6 levels at ICU admission and one week later. Compared to patients with CC/CT variants not treated with steroids, patients with the same genotype but receiving dexamethasone and patients with the TT variant not receiving steroids showed lower levels of IL-6 at admission (Figure 6E). After one week (Figure 6F), serum IL-6 levels remained lower in patients receiving dexamethasone with the CC/CT variant and in those with the TT variant and not treated with this drug. However, IL-6 levels were higher in those with the TT variant and treated with steroids, in line with our *ex-vivo* findings.

To explore the mechanisms behind these differences, peripheral blood gene expression at ICU day 4 was compared between patients treated with or without steroids for each rs1990760 variant. *IFIH1* expression decreased in patients with rs1990760 CC/CT variants treated with steroids, but not in those with a TT variant (Figure 6G). In opposite, steroids increased *FOXO3* expression only in patients with a TT variant (Figure 6H), resembling the results from the *ex-vivo* experiments. *IL6* gene raw counts at day 4 were below 5 in all the patients, and thus not compared. When the whole transcriptomes were compared, steroids significantly changed the expression of 58 genes in patients with a CC/CT variant and 23 in patients with a TT variant. Overall, changes in gene expression were qualitatively different between variants, with only three genes in common (Figure 6I and Supplementary Figure E6).

### In-silico clinical trials

These findings may support the hypothesis that COVID-19 populations with a low proportion of the rs1990760 TT variant would show a better response to dexamethasone. To explore this prediction, data from the RECOVERY trial (15) were examined. The T-allele frequency in populations from a Black/Asian ancestry is 0.13 (16), so only 1.7% has a TT variant. In these populations, RR was 0.7 (0.51-0.95), whereas in patients from a White ancestry (with a T allele frequency of 0.61), RR increased to 0.9 (0.8-1.2).

Using these data, RRs related to each rs1990760 variant, with and without steroids, were extracted and a survival model developed (see online supplement for details). A simulation including 500 patients from each rs1990760 variant and treatment arm (placebo or dexamethasone) showed a significant reduction in mortality with steroids in patients with a CC/CT variant, but not in those with a TT variant (Figure 7A). In patients not-receiving steroids, HRs related to a TT variant, compared to CC/CT variants, were independent of the allelic frequency (Figure 7B). Simulations of steroid therapy with different allelic frequencies showed that HRs in each specific variant remained constant whereas the effect of steroids in the overall population was dependent on the allele distribution (Figure 7C and Supplementary Figure E7).

**Figure 7.**
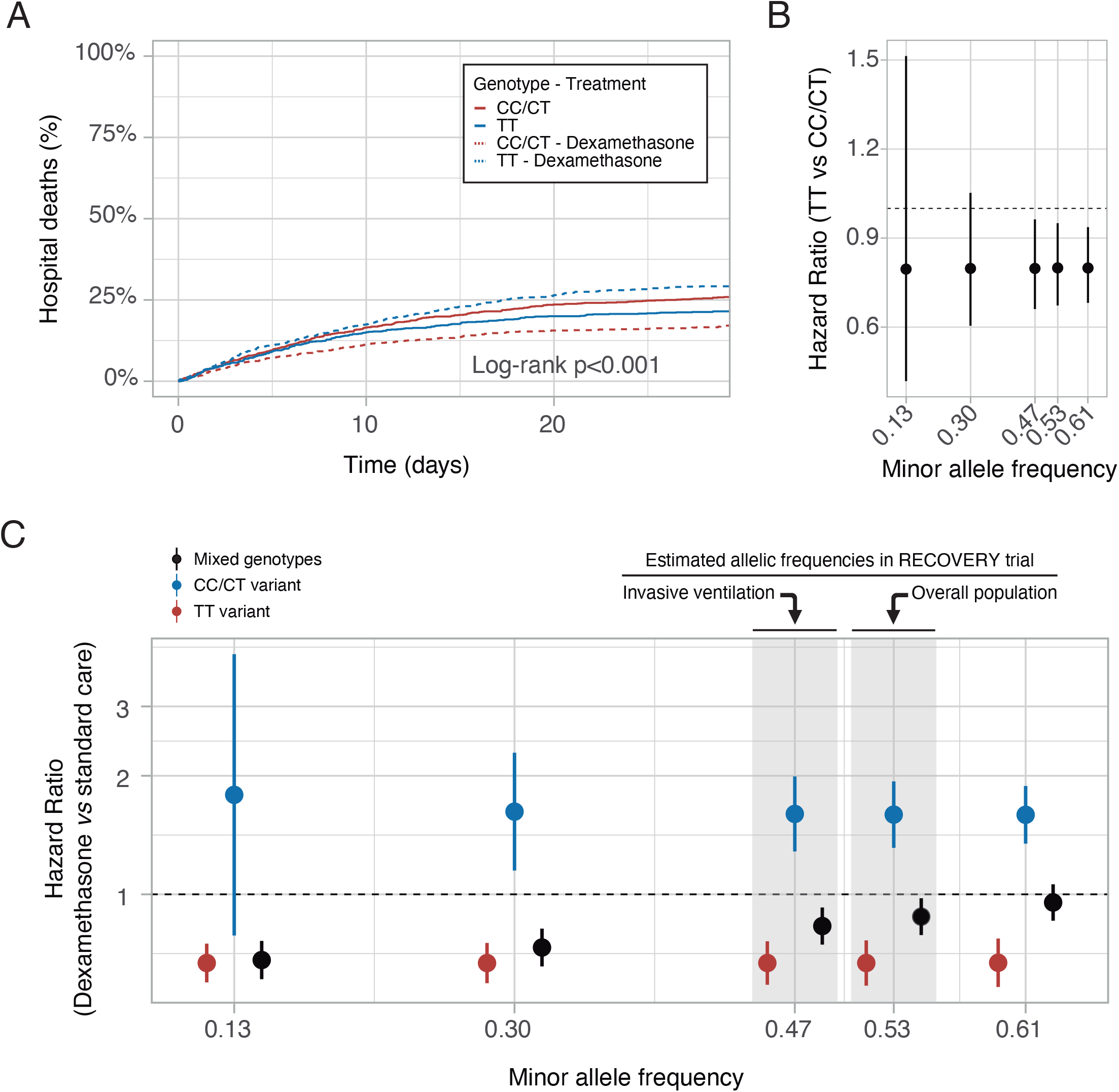
*In-silico* clinical trials. A: Mortality curves modelled using data from the RECOVERY clinical trial, for each rs1990760 variant and treatment allocation, assuming a sample size of 500 patients per group. B: Effect of rs1990760 TT variants (expressed as hazard ratio, HR) in simulated clinical trials including 6000 patients from populations with different minor allele frequencies, assigned to standard care. C: Effect of dexamethasone therapy according to rs1990760 variants and different minor allele frequencies in simulated clinical trials including 6000 patients. Minor allele frequencies were estimated according to reported race in several subsets of patients from the RECOVERY trial: European population (0.61), overall population (0.53), mechanically ventilated patients (0.47) and African American/Asian population (0.13). An intermediate value of 0.30 was added for illustrative purposes.

## Discussion

Our results show that critically ill patients with the TT variant in the *IFIH1* rs1990760 polymorphism have an attenuated inflammatory response to severe SARS-CoV-2 infection, leading to a decreased mortality. In this selected population, treatment with steroids has no immunomodulatory effects and could be related to worse outcomes. These results confirm the impact of the host response on patients’ outcomes and suggest that patient geno/phenotypes should be taken into account to prescribe steroids in this setting.

Host response is a major determinant of outcome in critically ill patients, including those with COVID-19. Several genetic variants involved in the inflammatory response have been related to SARS-CoV-2 infection and its severity (17–19). Activation of immune responses causes local and systemic inflammation, aimed to block viral replication. However, exacerbation of these responses can cause organ damage even after virus clearance. Our results recapitulate previous findings in COVID-19 describing the release of proinflammatory mediators, NK cell exhaustion, monocyte dysregulation and emergency hematopoiesis (20–22). Some of our findings in patients with the rs1990760 TT variant, including lower levels of circulating proinflammatory molecules and a shift towards anti-inflammatory cell populations (hematopoietic precursors, M2 macrophages or CD56^dim^ NK cells) have been linked to better outcomes in other observational studies (23). Collectively, our findings raise the hypothesis that the TT variant could be related to a lower *IFIH1* expression and an attenuated proinflammatory response. In line with this observation, a TT variant has been associated to lower proinflammatory cytokine levels in patients with lupus (24, 25) and in experimental viral infections (6).

Development of tolerance to viral diseases can be considered a major evolutionary adaptative response (26), and inhibition of pro-inflammatory responses has been proposed to improve the outcome of severe COVID-19 patients. Notably, pangolins, an intermediate host of coronaviruses (27), lack functional *IFIH1*. It has been suggested that this deficiency reduces the inflammatory response to coronavirus infections (28). IL-6 blockade (4) or steroids (3) are the only treatments that have improved the outcome of critically ill COVID-19 patients. However, mortality rates are still around 27-32% (3, 4), so new therapeutic approaches are warranted.

According to our results, the impact of the treatment with steroids along a given population may depend on intrinsic individual characteristics, allowing a personalized approach based on a specific genomic biomarker. In our study sample, patients with the *IFIH1* rs1990760 TT variant constitute a population with a better prognosis, in whom treatment with dexamethasone may be reconsidered as it was associated to higher mortality rates. Indeed, as shown by the *in-silico* trials, any population enriched for patients with CC/CT variants in rs1990760 will show higher mortality rates and a better response to steroids, due to the low proportion of TT variants. Dexamethasone in patients receiving mechanical ventilation at inclusion in the RECOVERY trial had a HR of 0.64, compared to a HR of 0.83 in the whole population. Of note, the proportion of Black/Asian patients (with a low frequency of a T allele) in the mechanically ventilated group was 29%, but only 18% in the whole study population. To date, no other genomic markers of personalized therapies in critically ill patients have been identified. The absence of differences in viral clearance and the late increase in IL-6 in these patients suggest that this worse outcome is more related to the disruption of ongoing immunoregulatory mechanisms than to antiviral responses. Our *ex-vivo* experiments do not fully elucidate the molecular mechanisms behind the interaction between TT variants and steroids, as the effects of *FOXO3* upregulation and *IL6* expression may be variable (29), but clearly illustrate that this specific combination may disrupt the ongoing self-regulation of inflammation. Moreover, steroid-induced changes in gene expression are qualitatively different in patients of each variant.

Our findings point towards the therapeutic potential of MDA5 modulation in COVID-19, either induced by steroids or targeted by other drugs. Amelioration of MDA5-dependent RNA sensing could avoid an exacerbated inflammatory response without impairing viral clearance. In this sense, it has been described that the interferon response triggered by MDA5 is unable to control viral replication (30). However, genetic ablation of *IFIH1* results in increased viral loads and decreased cytokine production (31), so this approach must be viewed with caution.

It is unclear if these findings can be translated to other viral diseases. Steroids may decrease mortality in an unselected ARDS population (32), but the role of underlying genotypes has not been addressed. An experimental model of Coxsackie virus infection revealed lower *IFIH1* and *CXCL10* expression in cells with the TT variant (6), with no relevant differences in viral clearance. However, total absence of MDA5 results in impaired clearance of West Nile virus (33).

Our work has several limitations. First, the results must be validated in an independent cohort. Our *in-silico* simulations reinforce the external validity of our findings, but a pharmacogenomic analysis of patients included in clinical trials is warranted for confirmation. Second, steroid treatment was not randomized, so we cannot discard other underlying factors responsible for the observed differences. Although there could be confounding by indication, there were no baseline differences among groups suggesting a higher severity in steroid-treated patients. Moreover, there are significant differences between genotypes irrespective of the treatment received. In addition, *in-silico* and *ex-vivo* experiments are congruent with the observed clinical results, supporting the differential effects of steroid therapy in *IFIH1* rs1990760 variants. Third, the favourable outcome of patients with a TT allele without steroids is based on a small sample size. As steroids are now the standard of care for severe COVID-19, this sample size cannot be increased outside a hypothetical clinical trial focused of personalized steroid therapy according to *IFIH1* rs1990760 variants. However, similar reduced numbers have served to identify other variants in the immune response (34), and the related finding of increased mortality in this genotype after steroid therapy compared to all other variants is supported by a larger sample. In addition, our sample is representative of European population with a limited racial diversity that may have influenced our results, as the *in-silico* analyses further suggest. Finally, other studies (35, 36) have focused on the genetic variants linked to an increased risk of severe COVID-19, compared to non-infected populations. However, no genetic markers have been associated to mortality in cohorts of infected patients. According to our data, rs1990760 is linked to the outcome, but no inferences on susceptibility to severe COVID-19 can be extracted.

In summary, we have identified a genetic variant of *IFIH1* that results in decreased expression of this gene and an ameliorated inflammatory response after severe SARS-CoV-2 infection. Patients with the rs1990760 TT genotype show a good outcome. However, this adaptative response was not observed in patients with a TT variant and treated with steroids. These findings suggest that the systemic response to severe COVID-19 is regulated by genetic factors that modulate the response to the infection and the prescribed therapy and, ultimately, may impact the outcome.

## Supporting information

Supplementary methods and results

## Data Availability

RNA-seq data has been deposited in Gene Expression Omnibus (accession numbers GSE168400 and GSE177025). Code for in-silico clinical trials is available at https://github.com/Crit-Lab/IFIH1_simulation. De-identified clinical data will be shared with researchers with any proposal after review and approval by the local Ethics Committee and a signed data access agreement. R code used for statistical analyses is also available on request. All requests should be sent to the corresponding author.

https://www.ncbi.nlm.nih.gov/geo/query/acc.cgi?acc=GSE168400

https://www.ncbi.nlm.nih.gov/geo/query/acc.cgi?acc=GSE177025

## Acknowledgements

The authors want to thank all the personnel at the participating ICUs and laboratories for their support during the development of the study. In addition, they thank Salvador Villalgordo, Silvia Viñas, Salvador Balboa, Rodrigo Albillos, Cecilia del Busto, Emilio García-Prieto, Jose Antonio Gonzalo, Diego Parra, Lisardo Iglesias, José Alonso, Sérida Dominguez and Alfredo González their additional efforts for sample collection.

## Data sharing

RNA-seq data has been deposited in Gene Expression Omnibus (accession numbers GSE168400 and GSE177025). Code for *in-silico* clinical trials is available at https://github.com/Crit-Lab/IFIH1_simulation. De-identified clinical data will be shared with researchers with any proposal after review and approval by the local Ethics Committee and a signed data access agreement. R code used for statistical analyses is also available on request. All requests should be sent to the corresponding author.

## References

1. Grasselli G, Zangrillo A, Zanella A, Antonelli M, Cabrini L, Castelli A, Cereda D, Coluccello A, Foti G, Fumagalli R, Iotti G, Latronico N, Lorini L, Merler S, Natalini G, Piatti A, Ranieri MV, Scandroglio AM, Storti E, Cecconi M, Pesenti A, COVID-19 Lombardy ICU Network. Baseline Characteristics and Outcomes of 1591 Patients Infected With SARS-CoV-2 Admitted to ICUs of the Lombardy Region, Italy. JAMA 2020;323:1574–1581.

2. Du Y, Tu L, Zhu P, Mu M, Wang R, Yang P, Wang X, Hu C, Ping R, Hu P, Li T, Cao F, Chang C, Hu Q, Jin Y, Xu G. Clinical Features of 85 Fatal Cases of COVID-19 from Wuhan. A Retrospective Observational Study. Am J Respir Crit Care Med 2020;201:1372–1379.

3. WHO Rapid Evidence Appraisal for COVID-19 Therapies (REACT) Working Group, Sterne JAC, Murthy S, Diaz JV, Slutsky AS, Villar J, Angus DC, Annane D, Azevedo LCP, Berwanger O, Cavalcanti AB, Dequin P-F, Du B, Emberson J, Fisher D, Giraudeau B, Gordon AC, Granholm A, Green C, Haynes R, Heming N, Higgins JPT, Horby P, Jüni P, Landray MJ, Le Gouge A, Leclerc M, Lim WS, Machado FR, et al. Association Between Administration of Systemic Corticosteroids and Mortality Among Critically Ill Patients With COVID-19: A Meta-analysis. JAMA 2020;324:1330–1341.

4. REMAP-CAP Investigators, Gordon AC, Mouncey PR, Al-Beidh F, Rowan KM, Nichol AD, Arabi YM, Annane D, Beane A, van Bentum-Puijk W, Berry LR, Bhimani Z, Bonten MJM, Bradbury CA, Brunkhorst FM, Buzgau A, Cheng AC, Detry MA, Duffy EJ, Estcourt LJ, Fitzgerald M, Goossens H, Haniffa R, Higgins AM, Hills TE, Horvat CM, Lamontagne F, Lawler PR, Leavis HL, et al. Interleukin-6 Receptor Antagonists in Critically Ill Patients with Covid-19. N Engl J Med 2021; doi:10.1056/NEJMoa2100433.

5. Gorman JA, Hundhausen C, Errett JS, Stone AE, Allenspach EJ, Ge Y, Arkatkar T, Clough C, Dai X, Khim S, Pestal K, Liggitt D, Cerosaletti K, Stetson DB, James RG, Oukka M, Concannon P, Gale M, Buckner JH, Rawlings DJ. The A946T variant of the RNA sensor IFIH1 mediates an interferon program that limits viral infection but increases the risk for autoimmunity. Nat Immunol 2017;18:744–752.

6. Domsgen E, Lind K, Kong L, Hühn MH, Rasool O, van Kuppeveld F, Korsgren O, Lahesmaa R, Flodström-Tullberg M. An IFIH1 gene polymorphism associated with risk for autoimmunity regulates canonical antiviral defence pathways in Coxsackievirus infected human pancreatic islets. Sci Rep 2016;6:39378.

7. Maiti AK. The African-American population with a low allele frequency of SNP rs1990760 (T allele) in IFIH1 predicts less IFN-beta expression and potential vulnerability to COVID-19 infection. Immunogenetics 2020;72:387–391.

8. Escudero D, Boga JA, Fernández J, Forcelledo L, Balboa S, Albillos R, Astola I, García-Prieto E, Álvarez-Argüelles ME, Martín G, Jiménez J, Vázquez F. SARS-CoV-2 analysis on environmental surfaces collected in an intensive care unit: keeping Ernest Shackleton’s spirit. Intensive Care Med Exp 2020;8:68.

9. Riviello ED, Kiviri W, Twagirumugabe T, Mueller A, Banner-Goodspeed VM, Officer L, Novack V, Mutumwinka M, Talmor DS, Fowler RA. Hospital Incidence and Outcomes of the Acute Respiratory Distress Syndrome Using the Kigali Modification of the Berlin Definition. Am J Respir Crit Care Med 2016;193:52–59.

10. Gómez-Novo M, Boga JA, Álvarez-Argüelles ME, Rojo-Alba S, Fernández A, Menéndez MJ, de Oña M, Melón S. Human respiratory syncytial virus load normalized by cell quantification as predictor of acute respiratory tract infection. J Med Virol 2018;90:861–866.

11. Patro R, Duggal G, Love MI, Irizarry RA, Kingsford C. Salmon provides fast and bias-aware quantification of transcript expression. Nat Methods 2017;14:417–419.

12. Love MI, Huber W, Anders S. Moderated estimation of fold change and dispersion for RNA-seq data with DESeq2. Genome Biol 2014;15:550.

13. Vallania F, Tam A, Lofgren S, Schaffert S, Azad TD, Bongen E, Haynes W, Alsup M, Alonso M, Davis M, Engleman E, Khatri P. Leveraging heterogeneity across multiple datasets increases cell-mixture deconvolution accuracy and reduces biological and technical biases. Nat Commun 2018;9:4735.

14. R Core Team. R: A language and environment for statistical computing. At <http://www.R-project.org/>.

15. RECOVERY Collaborative Group, Horby P, Lim WS, Emberson JR, Mafham M, Bell JL, Linsell L, Staplin N, Brightling C, Ustianowski A, Elmahi E, Prudon B, Green C, Felton T, Chadwick D, Rege K, Fegan C, Chappell LC, Faust SN, Jaki T, Jeffery K, Montgomery A, Rowan K, Juszczak E, Baillie JK, Haynes R, Landray MJ. Dexamethasone in Hospitalized Patients with Covid-19. N Engl J Med 2021;384:693–704.

16. COVID-19 GWAS Results. At <https://grasp.nhlbi.nih.gov/Covid19GWASResults.aspx>.

17. Bovijn J, Lindgren CM, Holmes MV. Genetic variants mimicking therapeutic inhibition of IL-6 receptor signaling and risk of COVID-19. Lancet Rheumatol 2020;2:e658–e659.

18. Zhang Q, Bastard P, Liu Z, Le Pen J, Moncada-Velez M, Chen J, Ogishi M, Sabli IKD, Hodeib S, Korol C, Rosain J, Bilguvar K, Ye J, Bolze A, Bigio B, Yang R, Arias AA, Zhou Q, Zhang Y, Onodi F, Korniotis S, Karpf L, Philippot Q, Chbihi M, Bonnet-Madin L, Dorgham K, Smith N, Schneider WM, Razooky BS, et al. Inborn errors of type I IFN immunity in patients with life-threatening COVID-19. Science 2020;370:eabd4570.

19. Gómez J, Albaiceta GM, Cuesta-Llavona E, García-Clemente M, López-Larrea C, Amado-Rodríguez L, López-Alonso I, Melón S, Alvarez-Argüelles ME, Gil-Peña H, Vidal-Castiñeira JR, Corte-Iglesias V, Saiz ML, Alvarez V, Coto E. The Interferon-induced transmembrane protein 3 gene (IFITM3) rs12252 C variant is associated with COVID-19. Cytokine 2021;137:155354.

20. Schulte-Schrepping J, Reusch N, Paclik D, Baßler K, Schlickeiser S, Zhang B, Krämer B, Krammer T, Brumhard S, Bonaguro L, De Domenico E, Wendisch D, Grasshoff M, Kapellos TS, Beckstette M, Pecht T, Saglam A, Dietrich O, Mei HE, Schulz AR, Conrad C, Kunkel D, Vafadarnejad E, Xu C-J, Horne A, Herbert M, Drews A, Thibeault C, Pfeiffer M, et al. Severe COVID-19 Is Marked by a Dysregulated Myeloid Cell Compartment. Cell 2020;182:1419–1440.e23.

21. Wilk AJ, Rustagi A, Zhao NQ, Roque J, Martínez-Colón GJ, McKechnie JL, Ivison GT, Ranganath T, Vergara R, Hollis T, Simpson LJ, Grant P, Subramanian A, Rogers AJ, Blish CA. A single-cell atlas of the peripheral immune response in patients with severe COVID-19. Nat Med 2020;26:1070–1076.

22. Wen W, Su W, Tang H, Le W, Zhang X, Zheng Y, Liu X, Xie L, Li J, Ye J, Dong L, Cui X, Miao Y, Wang D, Dong J, Xiao C, Chen W, Wang H. Immune cell profiling of COVID-19 patients in the recovery stage by single-cell sequencing. Cell Discov 2020;6:31.

23. Maucourant C, Filipovic I, Ponzetta A, Aleman S, Cornillet M, Hertwig L, Strunz B, Lentini A, Reinius B, Brownlie D, Cuapio A, Ask EH, Hull RM, Haroun-Izquierdo A, Schaffer M, Klingström J, Folkesson E, Buggert M, Sandberg JK, Eriksson LI, Rooyackers O, Ljunggren H-G, Malmberg K-J, Michaëlsson J, Marquardt N, Hammer Q, Strålin K, Björkström NK, Karolinska COVID-19 Study Group. Natural killer cell immunotypes related to COVID-19 disease severity. Sci Immunol 2020;5:eabd6832.

24. Zhang J, Liu X, Meng Y, Wu H, Wu Y, Yang B, Wang L. Autoimmune disease associated IFIH1 single nucleotide polymorphism related with IL-18 serum levels in Chinese systemic lupus erythematosus patients. Sci Rep 2018;8:9442.

25. Robinson T, Kariuki SN, Franek BS, Kumabe M, Kumar AA, Badaracco M, Mikolaitis RA, Guerrero G, Utset TO, Drevlow BE, Zaacks LS, Grober JS, Cohen LM, Kirou KA, Crow MK, Jolly M, Niewold TB. Autoimmune disease risk variant of IFIH1 is associated with increased sensitivity to IFN-α and serologic autoimmunity in lupus patients. J Immunol Baltim Md 1950 2011;187:1298–1303.

26. Schneider DS, Ayres JS. Two ways to survive infection: what resistance and tolerance can teach us about treating infectious diseases. Nat Rev Immunol 2008;8:889–895.

27. Xiao K, Zhai J, Feng Y, Zhou N, Zhang X, Zou J-J, Li N, Guo Y, Li X, Shen X, Zhang Z, Shu F, Huang W, Li Y, Zhang Z, Chen R-A, Wu Y-J, Peng S-M, Huang M, Xie W-J, Cai Q-H, Hou F-H, Chen W, Xiao L, Shen Y. Isolation of SARS-CoV-2-related coronavirus from Malayan pangolins. Nature 2020;583:286–289.

28. Fischer H, Tschachler E, Eckhart L. Pangolins Lack IFIH1/MDA5, a Cytoplasmic RNA Sensor That Initiates Innate Immune Defense Upon Coronavirus Infection. Front Immunol 2020;11:939.

29. Joseph J, Ametepe ES, Haribabu N, Agbayani G, Krishnan L, Blais A, Sad S. Inhibition of ROS and upregulation of inflammatory cytokines by FoxO3a promotes survival against Salmonella typhimurium. Nat Commun 2016;7:12748.

30. Rebendenne A, Valadão ALC, Tauziet M, Maarifi G, Bonaventure B, McKellar J, Planès R, Nisole S, Arnaud-Arnould M, Moncorgé O, Goujon C. SARS-CoV-2 triggers an MDA-5-dependent interferon response which is unable to control replication in lung epithelial cells. J Virol 2021;doi:10.1128/JVI.02415-20.

31. Yin X, Riva L, Pu Y, Martin-Sancho L, Kanamune J, Yamamoto Y, Sakai K, Gotoh S, Miorin L, De Jesus PD, Yang C-C, Herbert KM, Yoh S, Hultquist JF, García-Sastre A, Chanda SK. MDA5 Governs the Innate Immune Response to SARS-CoV-2 in Lung Epithelial Cells. Cell Rep 2021;34:108628.

32. Villar J, Ferrando C, Martínez D, Ambrós A, Muñoz T, Soler JA, Aguilar G, Alba F, González-Higueras E, Conesa LA, Martín-Rodríguez C, Díaz-Domínguez FJ, Serna-Grande P, Rivas R, Ferreres J, Belda J, Capilla L, Tallet A, Añón JM, Fernández RL, González-Martín JM, dexamethasone in ARDS network. Dexamethasone treatment for the acute respiratory distress syndrome: a multicentre, randomised controlled trial. Lancet Respir Med 2020;8:267–276.

33. Errett JS, Suthar MS, McMillan A, Diamond MS, Gale M. The essential, nonredundant roles of RIG-I and MDA5 in detecting and controlling West Nile virus infection. J Virol 2013;87:11416–11425.

34. Bastard P, Rosen LB, Zhang Q, Michailidis E, Hoffmann H-H, Zhang Y, Dorgham K, Philippot Q, Rosain J, Béziat V, Manry J, Shaw E, Haljasmägi L, Peterson P, Lorenzo L, Bizien L, Trouillet-Assant S, Dobbs K, de Jesus AA, Belot A, Kallaste A, Catherinot E, Tandjaoui-Lambiotte Y, Le Pen J, Kerner G, Bigio B, Seeleuthner Y, Yang R, Bolze A, et al. Autoantibodies against type I IFNs in patients with life-threatening COVID-19. Science 2020;370:eabd4585.

35. Pairo-Castineira E, Clohisey S, Klaric L, Bretherick AD, Rawlik K, Pasko D, Walker S, Parkinson N, Fourman MH, Russell CD, Furniss J, Richmond A, Gountouna E, Wrobel N, Harrison D, Wang B, Wu Y, Meynert A, Griffiths F, Oosthuyzen W, Kousathanas A, Moutsianas L, Yang Z, Zhai R, Zheng C, Grimes G, Beale R, Millar J, Shih B, et al. Genetic mechanisms of critical illness in Covid-19. Nature 2020;doi:10.1038/s41586-020-03065-y.

36. Severe Covid-19 GWAS Group, Ellinghaus D, Degenhardt F, Bujanda L, Buti M, Albillos A, Invernizzi P, Fernández J, Prati D, Baselli G, Asselta R, Grimsrud MM, Milani C, Aziz F, Kässens J, May S, Wendorff M, Wienbrandt L, Uellendahl-Werth F, Zheng T, Yi X, de Pablo R, Chercoles AG, Palom A, Garcia-Fernandez A-E, Rodriguez-Frias F, Zanella A, Bandera A, Protti A, et al. Genomewide Association Study of Severe Covid-19 with Respiratory Failure. N Engl J Med 2020;383:1522–1534.

