## Supplementary methods and results for "*IFIH1* rs1990760 variants, systemic inflammation and outcome in critically-ill COVID-19 patients"

**Online data supplement**

### Supplementary methods

*SARS-CoV-2 detection and quantification*

The presence of SARS-CoV-2 was analyzed by detecting viral genome using a multiple quantitative retrotranscriptase (RT)-PCR. Nucleic acids were purified by MagNa Pure 96 System (Roche, Geneva, Switzerland) from the swabs transport medium. The extracts were subjected to an amplification reaction using TaqMan Fast 1-Step Master Mix (Life technologies, Carlsbad, CA) supplemented with a mixture of primers (Thermo Fisher Scientific, Walthman, MA) and taqman MGB probes (Applied Biosystems, Foster City, CA) directed against ORF1ab and N genes (Table 1). Amplifications and subsequent analysis were carried out using the Applied Biosystems 7500 Real-time PCR System (1). Amplification of viral genes with a Ct number lower than 35 were considered positive. Viral load was normalized by the number of cells and expressed as copies/1000 cells, as previously described (2). Clearance of SARS-CoV-2 was evaluated by fitting viral load in tracheobronchial samples after its peak value over time using an exponential decay function and calculation of viral clearance half-life ($\lambda_{\text{Viral Clearance}}$).

*Genotyping*

DNA was extracted from total blood leukocytes (1 mL) with an automated equipment (Promega Maxwell). *IFIH1* rs1990760 C/T polymorphism determined by real time PCR with Taqman genotyping master mix (Life technologies) and Taqman probes (Thermo Fisher Scientific, assay C_2780299_30) in an ABI-7500 device. The genotyping strategy was validated by Sanger sequencing of selected individuals from the three genotypes.

*Blood sampling*

Two blood samples were taken in the first 24 hours after ICU admission. No patient had received steroids at the time of sampling. Three milliliters of blood were collected in Tempus tubes (Thermo Fisher) for RNA isolation and immediately stored at -80°C. Six additional milliliters were collected in a Vacutainer serum tube (BD Biosciences), and isolated by centrifugation and stored at -80°C until analysis.

*RNAseq*

After thawing, blood from Tempus tubes was diluted in PBS (1:3 v/v) and centrifuged at 3000 rpm for 30 min. Supernatant was discarded and the pellet resuspended in Trizol® (Sigma, Poole, UK) and precipitated overnight with isopropanol at -20°C. After centrifugation, RNA pellets were washed with 70% Ethanol and resuspended in RNAse-free water. RNA quality was assessed using a TapeStation, and only samples with a RIN (RNA integrity number) above 8 were analyzed.

RNA sequencing was performed using Ion AmpliSeq™ Transcriptome Human Gene Expression Kit, in an Ion S5 GeneStudio sequencer (Ion Torrent). Briefly, 10 ng of total RNA were retrotranscripted and the obtained cDNA used for library synthesis using Ion AmpliSeq™ Transcriptome kits to amplify all the canonical human transcripts. After template preparation in an automated Ion Chef Instrument, semiconductor 540 chips were run in an Ion S5 GeneStudio sequencer. Torrent Suite software was used for base calling, alignment and sequence quality controls. The generated FASTQ files (available at Gene Expression Omnibus, accession numbers GSE168400 and GSE 177025) were mapped against a reference transcriptome (obtained from http://refgenomes.databio.org) and transcripts counted using Salmon v1.4 software (3).

Raw counts were compared between genotypes using the DESeq2 library (4). The $\log_2$-fold change between variants for each gene and the adjusted p-value (corrected using a false discovery rate of 0.05) were calculated and analyzed using Ingenuity Pathway Analysis (Qiagen, USA) to identify overrepresented gene sets and networks. Over the identified network, *in-silico* effects of *IFIH1* down-regulation and addition of exogenous dexamethasone were performed.

*Peripheral blood cell populations*

Circulating cell populations were estimated from gene expression using a previously validated deconvolution algorithm (5). Using a reference expression matrix, proportions of 20 different cell lines were calculated. Only cell lines present in more than 5 samples were considered. As a quality check, we analyzed the correlation between estimated and measured lymphocyte percentages. The obtained correlation coefficient was 0.61 (Supplementary Figure E1). It must be noted, however, that the deconvolution method estimates proportions over transcriptionally active cells, which may not be equivalent to the obtained cell count (as there may be inactive cells in the latter).

*Inflammatory mediators*

A panel of inflammatory mediators was studied in serum from patients not receiving steroids during their first ICU day. Serum concentrations of interferons (IFN)-$\beta$, -$\gamma$ and -$\lambda$, tumor necrosis factor (TNF)-$\alpha$, interleukins (IL)-1$\beta$ and -6, and chemokines CXCL8, CXCL9, CXCL10, CXCL16, CCL2, CCL3, CCL4 and CCL7 were measured using a multiplexed assay (Luminex custom panel). Concentrations below the lower limit of detection for a given mediator were considered to be 0.

*Ex-vivo experiments*

To study the potential interferences between *IFIH1* rs1990760 variants and dexamethasone predicted by *in-silico* analyses, an ex-vivo experiment was designed. Blood samples from healthy volunteers (genotyped for *IFIH1* rs1990760 variants using DNA obtained from buccal swabs) were collected in EDTA tubes and immediately processed. Peripheral Blood Mononuclear Cells (PBMC) were isolated via density-gradient centrifugation with Lymphoprep (Axis-Shield PoC AS, Oslo, Norway). Cells were washed with red blood cell lysis buffer ($NH_4Cl$ 0.1M, $KHCO_3$ 0.01M, EDTA 0.1mM in $dH_2O$, pH 7.33) and PBS before being resuspended in RPMI-1640 (Gibco, USA) + 10% Fetal Bovine Serum (FBS). Cells were seeded in 12-well plates at a final concentration of $1{,}5 \times 10^6$ PBMC/ml and cultured at 37°C and 5% $CO_2$ in presence of medium/FBS, medium/FBS plus a MDA5 ligand (1 µg/ml high-molecular weight poly-I:C/LyoVec, Invivogen, USA), or medium plus the MDA5 ligand and 10µM dexamethasone (Kern Pharma, Spain). After 24h cells were collected and homogeneized in Trizol® for RNA extraction. 500 ng of total RNA was retrotranscribed into cDNA using an RT-PCR kit (High-capacity cDNA rt Kit, Applied Biosystems, USA). Expression of *STAT1*, *STAT3*, *FOXO3,* I*L6* and *GAPDH* was quantified using 5 ng of cDNA per well and in triplicate for each sample. Sybr-green Power up (Thermo Fisher Scientific) and 10 µM of the corresponding primers (Supplementary Table 1) were used in all the experiments. The relative expression of each gene was calculated as $2^{-\Delta CT(\text{gene of interest}) - \Delta CT(GAPDH)}$.

*Sample size calculation*

Given the exploratory nature of the study objective and the absence of previous data, no formal sample size calculations were performed. The study started after approval from the ethics committee and finished in December 2020, after the second pandemic wave.

*Statistical analysis*

Data are shown as median (interquartile range). Comparisons between *IFIH1* variants were done using Wilcoxon or ANOVA tests, and p-values corrected using a false discovery rate of 0.05. Results from the *ex-vivo* model were fitted to a mixed effects model including experimental group and genotype as covariables, and post-hoc comparisons evaluated using Holm's correction.

Survival was analyzed using a competing risks, Cox regression model, with ICU/hospital discharge alive and spontaneously breathing and death as competing risks, using the Aalen-Johansen estimator, as previously described (6). This competing-risks framework is needed as patients discharged alive have a low probability of death, so censoring at the time of discharge using a standard Kaplan-Meier approach would lead to biased observation, as the probability of death is different as those still followed (i.e. informative censoring). Patients with a rs1990760 CC/CT variant not treated with steroids were considered the reference category in all the analyses.

All the analyses and plots were performed using the R 4.0.1 statistical environment (7) with the packages data.table (8), multcomp (9), survival (10), MetaIntegrator (11) and ggplot2 (12).

*Estimation of Hazard Ratios from the RECOVERY trial*

Our findings raise the hypothesis that steroid therapy will have a larger effect in populations with a low proportion of individuals with a TT genotype. Populations with black/asian ancestry have an allelic frequency of the T allele of 0.13 (13). Therefore, the frequency of a TT genotype is 0.017, implying that the majority of the population has a CC/CT variant. Using data from the RECOVERY trial (14), we assume that the effects of steroids in this population correspond to the effect on patients with a CC/CT variant. In this population, steroids significantly decreased mortality, with a risk ratio (RR) of 0.7 (0.51-0.95). Therefore, we assumed that treatment with steroids in patients with these non-TT variants decreases mortality with a RR of 0.7.

White populations have an allelic frequency of the T allele in rs1990760 of 0.61 (13). Therefore, the proportion of patients with a TT variant is 0.37. GWAS data released by the UK biobank report a coefficient for hospital death related to the T allele of 0.03 (13), so the RR for a TT variant (irrespective of the therapy) is $e^{2\cdot 0.03}$ =1.06. Translating these findings to white patients included in the RECOVERY trial yields the following mortality table (assuming that randomization was independent of the rs1990760 variant):

| Genotype | Patients | Standard care | Dexamethasone |
|---|---|---|---|
| | 4689 | 849 / 3139 | 401 / 1550 |
| CC/CT | 2954 | a / 1978 | b / 976 |
| TT | 1735 | (849-a) /1161 | (401-b) / 574 |

Where a and b are the number of deaths in patients with a CC/CT variant assigned to standard care or dexamethasone, respectively.

Considering that the previously calculated RR of steroids in CC/CT population is 0.7, we can estimate:

$$\frac{b}{976} = 0.7\ \frac{a}{1978}$$

$$b = 0.3454\ a$$

And assuming a global RR of a TT genotype of 1.06 (irrespective of therapy), we can estimate:

$$\frac{\frac{(849 - a) + (401 - b)}{1735}}{\frac{a + b}{2954}} = 1.06$$

Solving these two equations yields the following values:

$$a = 573$$

$$b = 198$$

Therefore, distribution of mortality according to therapy and rs1990760 variants was estimated as:

| Genotype | Patients | Standard care | Dexamethasone |
|---|---|---|---|
| | 4689 | 849 / 3139 | 401 / 1550 |
| CC/CT | 2954 | 573 / 1978 | 198 / 976 |
| TT | 1735 | 276 /1161 | 203 / 574 |

From these values, the RR of a TT genotype compared to CC/CT variants in patients not receiving steroids is:

$$RR_{\left(TT\ vs \frac{CC}{CT}\right)\ No\ steroids} = \frac{\frac{276}{1161}}{\frac{573}{1978}} = \frac{0.238}{0.290} = 0.821$$

And the RR related to steroid therapy compared to standard care in patients with a TT variant is:

$$RR_{(Dexamethasone\ vs\ std.care)TT\ variant} = \frac{\frac{203}{574}}{\frac{276}{1161}} = \frac{0.354}{0.238} = 1.487$$

*In-silico clinical trial*

The previously estimated RRs were used to simulate mortality curves and perform *in-silico* clinical trials. Mortality was modelled using an asymptotic regression curve with a cumulative distribution function:

$$F(x) = a - ae^{-ct}$$

Where *a* is the upper asymptote, *c* is a curvature parameter, *e* is the base of natural logarithms and *t* is time. Assuming that final mortality (this is, the upper asymptote) is 10% higher than the observed mortality at day 28 (as in the survival curves of our cohort), we obtain that the curvature parameter is 0.08564. This parameter is constant for all curves, so the differences among groups depend only on the upper asymptote. This model is appropriate to fit acute conditions in which most of the deaths occur in the short term, but a substantial proportion of patients survive the disease. The code for these simulations can be found at *https://github.com/Crit-Lab/IFIH1_simulation*.

Different clinical trials including patients randomized into four groups depending on the rs1990760 variant (CC/CT or TT) and prescription of dexamethasone were simulated. Randomization to dexamethasone or placebo was 1:1. Minor allele frequencies from 0.13 to 0.61 (corresponding to populational distributions) and two baseline mortality rates (by adjusting the mortality of patients with a CC/CT variant not treated with steroids to 25.7% and 41.4%, corresponding to mortality rates of the overall RECOVERY trial and those receiving mechanical ventilation respectively) were tested. Each set of conditions was repeated 1000 times and the average RR calculated. All the simulations were done using the Mediana package (15) for R.

### Supplementary results

**Supplementary Table E1.** Primers and probes used to detect SARS-CoV-2.

| Design | Position | Name | Sequence (5´-3´) | Gen |
|---|---|---|---|---|
| In house | Sense primer | CoV-2-OVI-S | ATCAAGTTAATGGTTACCCTAACATGT | |
| | Antisense primer | CoV-2-OVI-A | AACCTAGCTGTAAAGGTAAATTGGTACC | ORF1ab |
| | Probe MGB FAM | CoV-2-OVI-FAM | CCGCGAAGAAGCTA | |
| CDC[1] | Sense primer | 2019-nCoV_N1-F | GACCCCAAAATCAGCGAAAT | |
| | Antisense primer | 2019-nCoV_N1-R | TCTGGTTACTGCCAGTTGAATCTG | Gen N |
| | Probe MGB VIC | 2019-nCoV_N1-P-VIC | CCGCATTACGTTTGGT[2] | |

[1] Sequences obtained from reference (16)

[2] Probe sequence has been shortened  as it is a MGB probe

**Supplementary Table E2.** Primers used for qPCR of human genes.

| Gene | Forward | Reverse |
| --- | --- | --- |
| ***STAT1*** | 5'-CCGTTTTCATGACCTCCTGT-3' | 5'-TGAATATTCCCCGACTGAGC-3' |
| ***STAT3*** | 5'-TTTCACTTGGGTGGAGAAGG-3' | 5'-GCTACCTGGGTCAGCTTCAG-3' |
| ***FOXO3*** | 5'-ACAAACGGCTCACTCTGTCC-3' | 5'-TCTTGCCAGTTCCCTCATTC-3' |
| ***IL6*** | 5'-GGTACATCCTCGACGGCATCT-3' | 5'-GTGCCTCTTTGCTGCTTTCAC-3' |
| ***GAPDH*** | 5'-TCGGAGTCAACGGATTTGGTCGT-3' | 5'-TGCCATGGGTGGAATCATATTGGA-3' |

**Supplementary Table E3.** Clinical characteristics of patients according to *IFIH1* rs1990760 genotypes and dexamethasone therapy. Values are shown as absolute count or median (interquartile range). PBW: Predicted body weight. NIV: Non-invasive ventilation. PEEP: Positive End-Expiratory Pressure. *p values calculated for proportion over the number of intubated patients.

| | No steroids | | Steroids | | |
|---|---|---|---|---|---|
| Rs1990760 ► | CC/CT | TT | CC/CT | TT | P value |
| Age (y) | 68 (59 - 73) | 64.5 (59 - 68) | 66 (57 - 75) | 70 (63 - 76) | 0.209 |
| Sex | | | | | 0.204 |
| Male | 30 | 8 | 97 | 39 | |
| Female | 5 | 6 | 30 | 12 | |
| Race | | | | | 0.746 |
| Black | 0 | 0 | 2 | 1 | |
| White | 34 | 13 | 112 | 48 | |
| Latino | 1 | 1 | 12 | 1 | |
| Asian | 0 | 0 | 1 | 0 | |
| Body mass index (Kg/m$^2$) | 28 (25 - 33) | 32 (29 - 34) | 30 (27 - 33) | 31 (26 - 34) | 0.566 |
| Days since symptom onset | 7 (7 – 10) | 10 (6 – 11) | 8 (6 – 11) | 8 (6 -10) | 0.857 |
| Days from hospital admission to ICU admission | 1 (0 - 3) | 1 (1 – 3) | 2 (1 – 4) | 2 (1 – 3) | 0.262 |
| Arterial hypertension | 20 | 10 | 74 | 28 | 0.77 |
| Diabetes | 9 | 4 | 28 | 10 | 0.87 |
| Chronic kidney disease | 4 | 0 | 7 | 5 | 0.577 |
| COPD | 3 | 1 | 7 | 5 | 0.735 |
| Cirrhosis | 0 | 0 | 1 | 1 | 0.761 |
| Neoplasms | | | | | 0.009 |
| No | 29 | 13 | 125 | 48 | |
| Active | 4 | 1 | 3 | 2 | |
| Past | 2 | 0 | 0 | 0 | |
| Ventilation at admission | | | | | 0.525 |
| Spontaneous / NIV | 6 | 4 | 21 | 7 | |
| Controlled invasive ventilation | 29 | 10 | 106 | 42 | |
| Pressure support ventilation | 0 | 0 | 0 | 1 | |
| $FiO_2$ | 0.5 (0.4 - 0.6) | 0.45 (0.4 - 0.6) | 0.5 (0.4 - 0.6) | 0.5 (0.4 - 0.6) | 0.069 |
| $PaO_2/FiO_2$ (mmHg) | 189 (148 - 237) | 180 (102 - 231) | 220 (171 - 297) | 188 (144 - 223) | 0.006 |
| $PaCO_2$ (mmHg) | 42 (39 - 47) | 43 (42 - 45) | 43 (39 - 48) | 44 (38 - 47) | 0.795 |
| Respiratory rate (min$^{-1}$) | 18 (16 - 20) | 19 (18 - 21) | 18 (16 - 22) | 18 (16 - 20) | 0.834 |
| Arterial pH | 7.36 | 7.39 | 7.37 | 7.38 | 0.712 |

| | | | | | |
|---|---|---|---|---|---|
| | (7.32 - 7.4) | (7.36 - 7.42) | (7.32 - 7.42) | (7.33 - 7.4) | |
| Tidal volume/PBW (ml/Kg) | 7.3 (6.9 - 8.1) | 8.4 (7.9 - 9.2) | 7.5 (6.9 - 8.2) | 7.8 (6.7 - 9.2) | 0.134 |
| Plateau pressure ($cmH_2O$) | 25 (22 - 29) | 27 (26 - 29) | 23 (21 - 27) | 24 (20 - 29) | 0.2 |
| PEEP ($cmH_2O$) | 14 (10 - 16) | 14 (12 - 14) | 12 (10 - 14) | 12 (10 - 14) | 0.417 |
| Driving pressure ($cmH_2O$) | 12 (10 - 14) | 14 (12 - 14) | 12 (10 - 14) | 12 (10 - 14) | 0.728 |
| Respiratory system compliance ($ml/cmH_2O$) | 39 (32 - 53) | 36 (31 - 43) | 42 (32 - 50) | 37 (31 - 46) | 0.471 |
| Leukocytes ($x10^3/\mu l$) | 8.20 (5.56 – 10.42) | 6.90 (5.94 – 9.68) | 8.33 (6.14 – 11.44) | 9.35 (6.01 – 11.68) | 0.836 |
| Lymphocytes ($x10^3/\mu l$) | 0.68 (0.59 – 0.90) | 0.79 (0.58 – 12.67) | 0.66 (0.48 – 0.94) | 0.57 (0.45 – 0.91) | 0.028 |
| Serum creatinine (mg/dl) | 0.92 (0.68 - 1.41) | 0.81 (0.58 - 1.05) | 0.77 (0.61 - 1.06) | 0.87 (0.64 - 1.24) | 0.104 |
| Serum ferritin (ng/ml) | 1582 (1062 - 2576) | 645 (327 - 1025) | 1125 (757 - 1896) | 1015 (712 - 1389) | 0.004 |
| D-dimer (ng/ml) | 1098 (776 - 2351) | 1095 (717 - 1522) | 1026 (659 - 2026) | 1085 (633 - 1806) | 0.513 |
| Vasoactive drugs | 26 | 9 | 61 | 28 | 0.042 |
| Invasive mechanical ventilation | 34 | 12 | 117 | 47 | 0.561 |
| Neuromuscular blocking agents | 19 | 3 | 50 | 27 | 0.097* |
| Prone ventilation | 24 | 4 | 63 | 28 | 0.117* |
| ECMO | 1 | 0 | 2 | 2 | 0.735* |
| ICU length of stay (days) | | | | | |
| Overall | 19 (13 - 33) | 13 (10 - 16) | 13 (9 - 31) | 15 (10 - 22) | 0.451 |
| Survivors | 15 (13 – 24) | 13 (10 – 16) | 12 (8 – 29) | 17 (8 – 22) | 0.620 |
| ICU mortality | 8 | 0 | 27 | 18 | 0.034 |
| Hospital mortality | 9 | 0 | 27 | 19 | 0.020 |

**Supplementary figure E1.** Correlation between measured and estimated (from deconvolution analysis) lymphocyte percentages in peripheral blood.

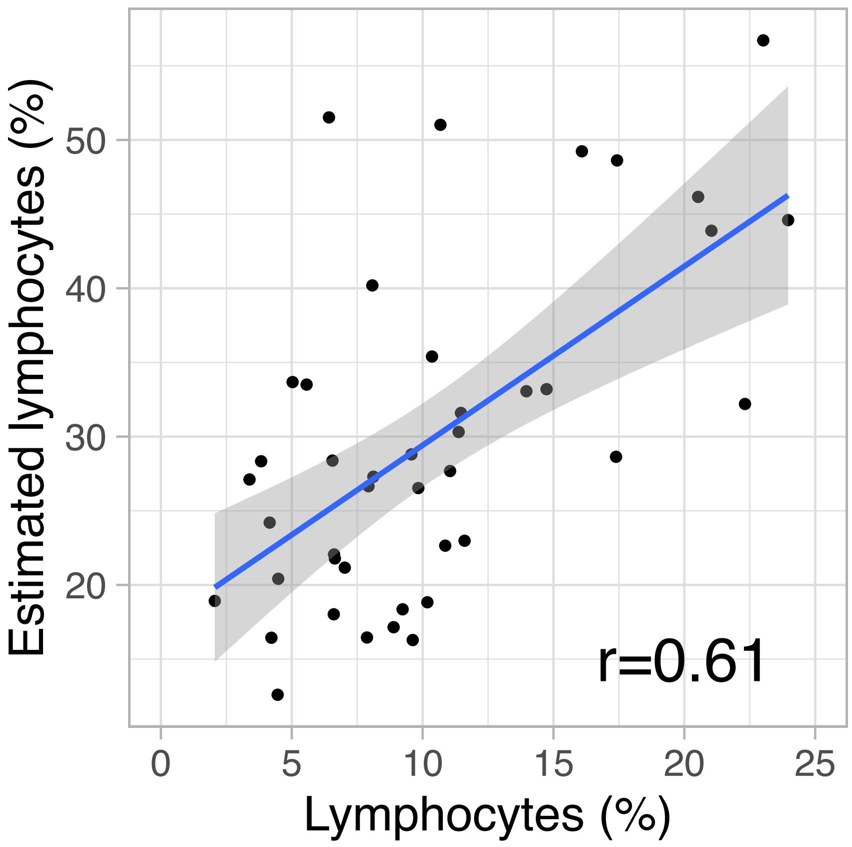

**Supplementary Figure E2.** *IFIH1* expression per rs1990760 variants.

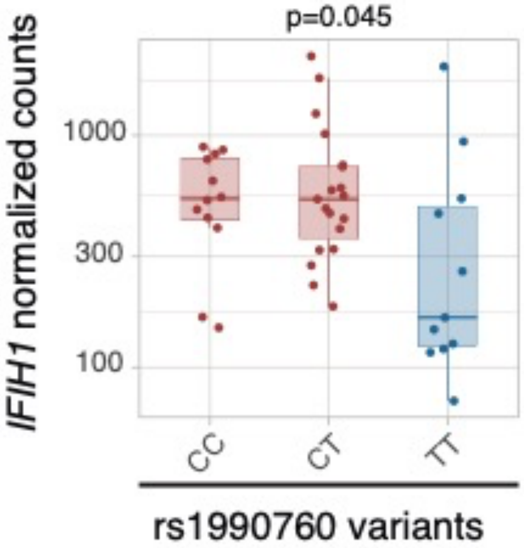

**Supplementary Figure E3.** Gene networks involved in regulation of the inflammatory response identified by Ingenuity Pathway Analysis among the genes with differential expression in patients with rs1990760 CC/CT and TT variants.

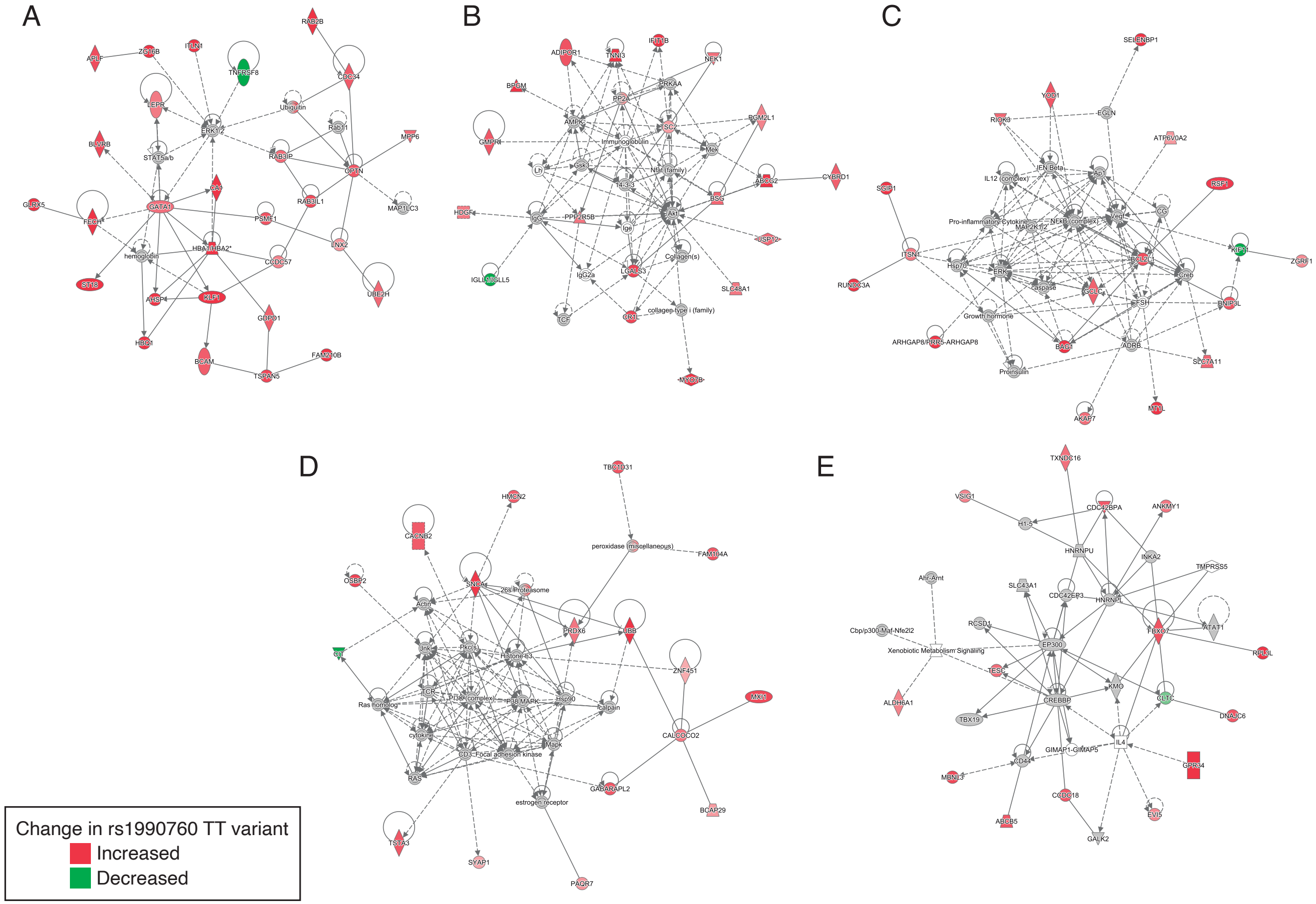

**Supplementary Figure E4.** *In-silico* predictions of *IFIH1* downregulation over a gene network involved in regulation of the inflammatory response.

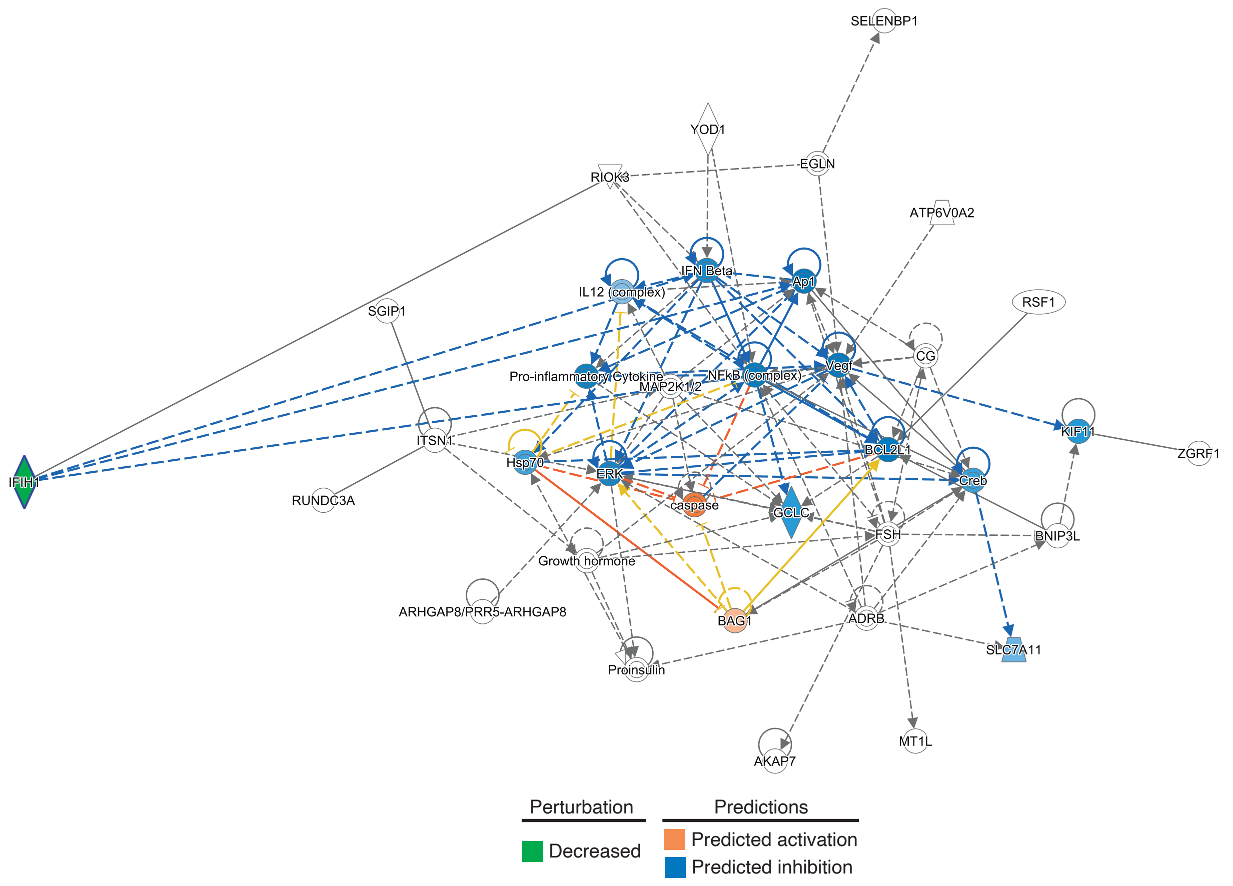

**Supplementary Figure E5-** Predicted effects of *IFIH1* downregulation in absence (A) or in presence (B) of dexamethasone. A number of molecules down-regulated in response to lower *IFIH1* expression are up-regulated when dexamethasone is added (arrowheads).

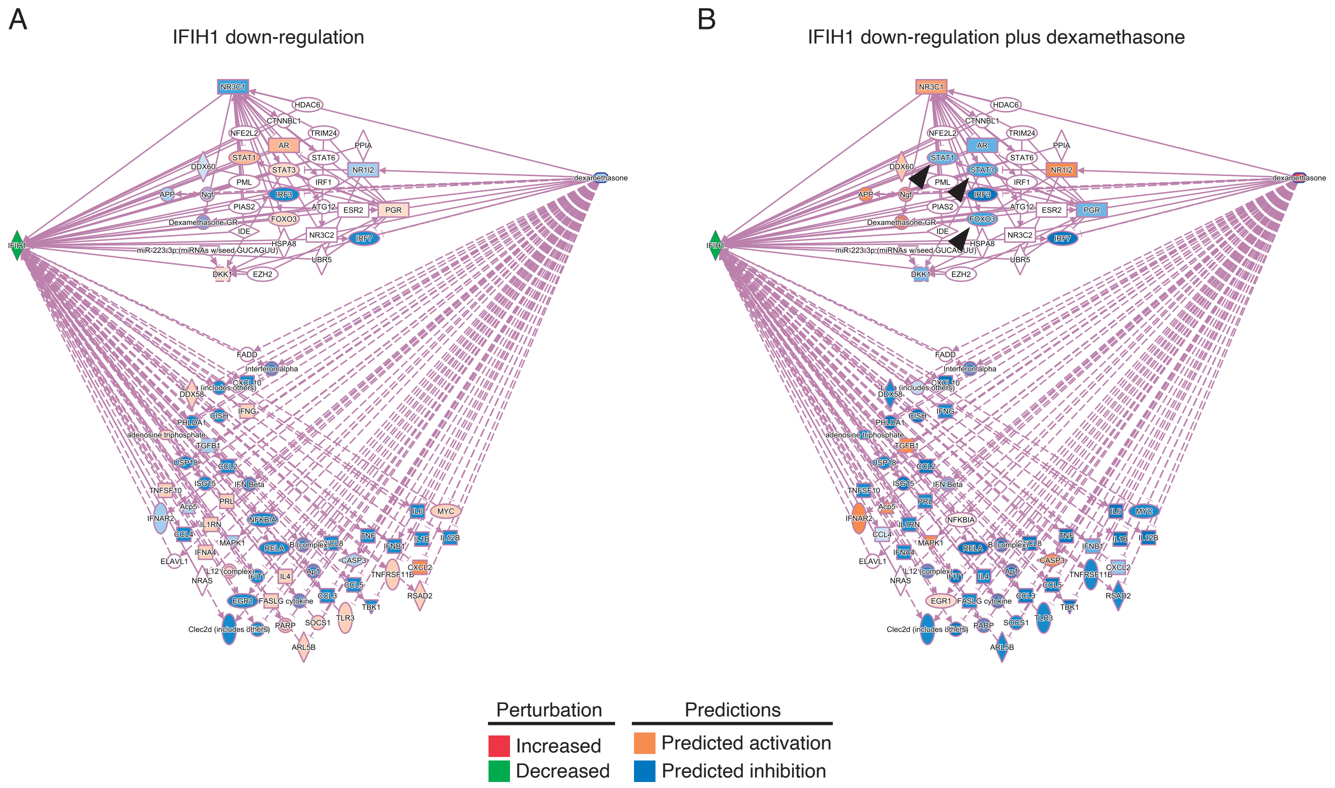

**Supplementary Figure E6.** Heatmaps with the differentially expressed genes in response to steroids in patients of each rs1990760 variant. Transcriptomic signatures in each variant only share 3 genes.

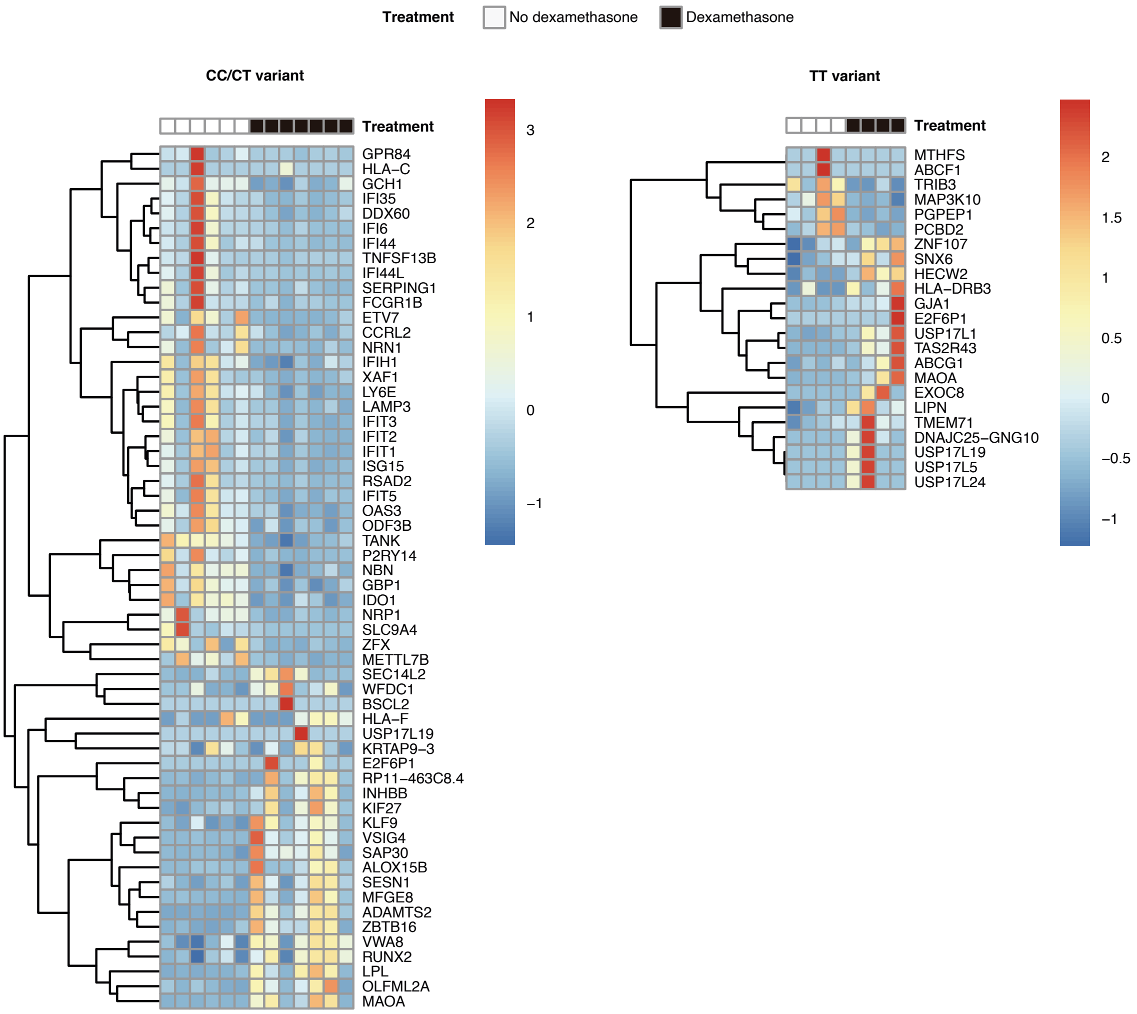

**Supplementary Figure E7.** Simulated survival curves corresponding to *in-silico* clinical trials testing dexamethasone in 6000 COVID-19 patients from populations with different allelic frequencies. T allele frequencies correspond to an African/American/Asian population (0.13), mechanically ventilated sample of the RECOVERY trial (0.47), overall sample of the RECOVERY trial (0.53) and a White population (0.61).

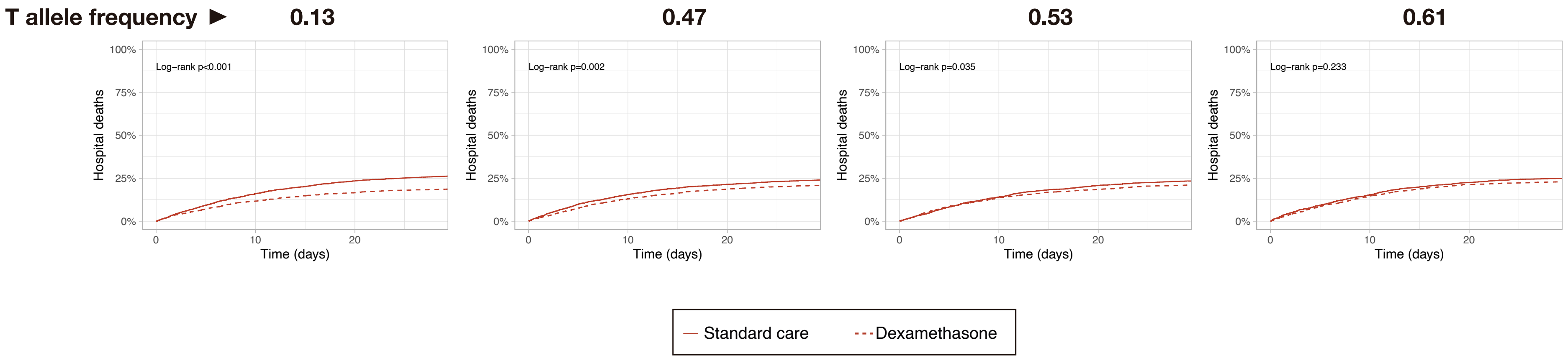

**Supplementary References**

1. Escudero D, Boga JA, Fernández J, Forcelledo L, Balboa S, Albillos R, Astola I, García-Prieto E, Álvarez-Argüelles ME, Martín G, Jiménez J, Vázquez F. SARS-CoV-2 analysis on environmental surfaces collected in an intensive care unit: keeping Ernest Shackleton's spirit. *Intensive Care Med Exp* 2020;8:68.
2. Gómez-Novo M, Boga JA, Álvarez-Argüelles ME, Rojo-Alba S, Fernández A, Menéndez MJ, de Oña M, Melón S. Human respiratory syncytial virus load normalized by cell quantification as predictor of acute respiratory tract infection. *J Med Virol* 2018;90:861–866.
3. Patro R, Duggal G, Love MI, Irizarry RA, Kingsford C. Salmon provides fast and bias-aware quantification of transcript expression. *Nat Methods* 2017;14:417–419.
4. Love MI, Huber W, Anders S. Moderated estimation of fold change and dispersion for RNA-seq data with DESeq2. *Genome Biol* 2014;15:550.
5. Vallania F, Tam A, Lofgren S, Schaffert S, Azad TD, Bongen E, Haynes W, Alsup M, Alonso M, Davis M, Engleman E, Khatri P. Leveraging heterogeneity across multiple datasets increases cell-mixture deconvolution accuracy and reduces biological and technical biases. *Nat Commun* 2018;9:4735.
6. Barbaro RP, MacLaren G, Boonstra PS, Iwashyna TJ, Slutsky AS, Fan E, Bartlett RH, Tonna JE, Hyslop R, Fanning JJ, Rycus PT, Hyer SJ, Anders MM, Agerstrand CL, Hryniewicz K, Diaz R, Lorusso R, Combes A, Brodie D, Extracorporeal Life Support Organization. Extracorporeal membrane oxygenation support in COVID-19: an international cohort study of the Extracorporeal Life Support Organization registry. *Lancet* 2020;396:1071–1078.
7. R Core Team. R: A language and environment for statistical computing. at <http://www.R-project.org/>.
8. Dowle M, Srinivasan A. *data.table: Extension of `data.frame`*. 2021. at <https://CRAN.R-project.org/package=data.table>.
9. Hothorn T, Bretz F, Westfall P. Simultaneous Inference in General Parametric Models. *Biom J* 2008;50:346–363.
10. Therneau TM. *A Package for Survival Analysis in R*. 2020. at <https://CRAN.R-project.org/package=survival>.
11. Haynes WA, Vallania F, Liu C, Bongen E, Tomczak A, Andres-Terrè M, Lofgren S, Tam A, Deisseroth CA, Li MD, Sweeney TE, Khatri P. Empowering multi-cohort gene expression analysis to increase reproducibility. *Pac Symp Biocomput Pac Symp Biocomput* 2017;22:144–153.
12. Wickham H. *ggplot2: Elegant Graphics for Data Analysis*. Springer-Verlag New York; 2016. at <https://ggplot2.tidyverse.org>.
13. COVID-19 GWAS Results. at <https://grasp.nhlbi.nih.gov/Covid19GWASResults.aspx>.
14. RECOVERY Collaborative Group, Horby P, Lim WS, Emberson JR, Mafham M, Bell JL, Linsell L, Staplin N, Brightling C, Ustianowski A, Elmahi E, Prudon B, Green C, Felton T, Chadwick D, Rege K, Fegan C, Chappell LC, Faust SN, Jaki T, Jeffery K, Montgomery A, Rowan K, Juszczak E, Baillie JK, Haynes R, Landray MJ. Dexamethasone in Hospitalized Patients with Covid-19. *N Engl J Med* 2021;384:693–704.
15. Paux G, Dmitrienko A. *Mediana: Clinical Trial Simulations*. 2019. at <https://CRAN.R-project.org/package=Mediana>.

16. US Centers for Disease Control and Prevention. 2019-Novel coronavirus (2019-nCoV) real-time rRT-PCR panel primers and probes. *Cent Dis Control Prev* 2020;at <https://www.cdc.gov/coronavirus/2019-ncov/lab/rt-pcr-panel-primer-probes.html>.
